# Impact of supplementary air filtration on airborne particulate matter in a UK hospital ward

**DOI:** 10.1101/2022.03.25.22272953

**Authors:** D Sloof, MB Butler, C Peters, A Conway Morris, T Gouliouris, R Thaxter, VL Keevil, CB Beggs

**Affiliations:** AirPurity UK, Ltd, Cambridge, UK; Department of Medicine for the Elderly, Cambridge University Hospitals, UK; Department of Microbiology, NHS Greater Glasgow and Clyde, UK; John V Farman Intensive Care Unit, Cambridge University Hospitals, UK; Division of Immunology, Department of Pathology, University of Cambridge, UK; Department of Medicine, University of Cambridge, UK; Infection Prevention and Control, Cambridge University Hospitals, UK; Carnegie School of Sport, Leeds Beckett University, UK

## Abstract

**Background:** During the COVID-19 pandemic, aerosol spread of SARS-CoV-2 has been a major problem in healthcare facilities, resulting in increased use of supplementary HEPA filtration to mitigate transmission. We report here a natural experiment that occurred when an air filtration unit (AFU) on an inpatient ward for older people was accidentally switched off.

**Aim:** To assess aerosol transport within the ward and determine whether the AFU reduced particulate matter (PM) levels in the air.

**Methods:** Time-series PM, CO_2_, temperature and humidity data (at 1 minute intervals) was collected from multiple sensors around the ward over two days in August 2021. During this period, the AFU was accidentally switched off for approximately 7 hours, allowing the impact of the intervention on particulates (PM1-PM10) to be assessed using a Mann-Whitney test. Pearson correlation analysis of the PM and CO_2_ signals was also undertaken to evaluate the movement of airborne particulates around the ward.

**Findings:** The AFU greatly reduced PM counts of all sizes throughout the ward space (p<0.001 for all sensors), with PM signals positively correlated with indoor CO_2_ levels (r = 0.343 – 0.817; all p<0.001). Aerosol particle counts tended to rise and fall simultaneously throughout the ward space when the AFU was off, with PM signals from multiple locations highly correlated (e.g. r = 0.343 – 0.868 (all p<0.001) for PM1).

**Conclusion:** Aerosols freely migrated between the various sub-compartments of the ward, suggesting that social distancing measures alone cannot prevent nosocomial transmission of SARS-CoV-2. The AFU greatly reduced PM levels throughout the ward space.

**Practical implications:**

- Aerosols can freely migrate throughout whole wards, suggesting that social distancing measures alone are not enough to prevent SARS-CoV-2 transmission.
- Appropriately sized supplementary room air filtration, if utilised correctly, can greatly reduce aerosol levels throughout ward spaces.
- Air filtration devices are often placed in rooms without any consideration given to their performance. It is therefore important to commission air filtration devices using PM and CO_2_ sensors before they are utilised in order to demonstrate that they are effective throughout entire ward spaces.

## Introduction

The COVID-19 pandemic has led to major advances in understanding how respiratory viral infections are spread within buildings, and it is now known that small aerosol particles play a dominant role in the transmission of SARS-CoV-2 [1-6]. These are formed when exhaled respiratory droplets, <100 µm in diameter, evaporate to become aerosol particles [7, 8] approximately 20–34% of their original size [9]. Particles of this size can remain suspended in the air for many minutes [10] and can be readily inhaled, with those in the size range 2.5 - 20 µm thought to account for 90% of the viral transmission at the nasopharynx [9]. As such, transmission of SARS-CoV-2 is thought to primarily occur when infectious aerosol particles in this size range come into contact with angiotensin-converting enzyme 2 (ACE2) receptors in the nasopharynx. [11]. However, ACE2 receptors are also found throughout the respiratory tree including on the alveolar epithelial cells, and therefore inhalation of smaller infectious aerosol particles <5 µm that can travel deeper into the lungs may also contribute to the burden of disease [12, 13].

Infectious respiratory aerosols can be liberated in large quantities when talking, singing, or simply breathing [14-16] and may build up to high concentrations in room air, if the space is not adequately ventilated [10]. Consequently, poorly ventilated spaces that may contain infectious individuals, such as hospital wards, can pose a considerable threat to patients and healthcare workers (HCWs) alike, with numerous nosocomial COVID-19 outbreaks reported in the literature [17-25]. The problem can be particularly acute in wards containing older and/or immunocompromised patients who are vulnerable to developing severe disease following viral infection. Furthermore, in open-plan wards with multi-bedded bays, pressure gradients may exist due to room mechanical ventilation or wind pressure, with the result that respiratory aerosol particles can migrate considerable distances. As such, vulnerable patients who are at some distance (>2 m) from an infector can become exposed [26]. Realisation of this issue has led to growing interest in non-pharmaceutical interventions such as supplementary room air filtration [27] and air disinfection [28, 29], and also utilising carbon dioxide (CO_2_) monitoring [30, 31] to optimise ventilation [32]. These aim to mitigate the transmission of SARS-CoV-2 in clinical settings, and it is within this context that we report our findings.

Here we report the results of a natural experiment that occurred on a medicine for older people ward at an NHS University hospital in the UK on the 3^rd^ and 4^th^ August 2021, when a room air filtration unit (AFU), containing high efficiency particulate air (HEPA) filters and an ultraviolet (UV) air disinfection lamp (at 254 nm), was being commissioned. The experiment arose because the AFU was accidentally switched off for several hours. We were able to analyse in detail the impact of this natural intervention on the airborne particulate matter (PM) and CO_2_ levels in the ward space, because the space was fitted with automatic sensors that recorded minute-by-minute PM and CO_2_ levels in the air, as well as temperature and relative humidity (RH). Comparing these markers of air quality across similar time periods on consecutive days, when the AFU was switched off and then turned back on again, we were able to test the hypothesis that PM levels in the air were higher when the AFU was not in operation compared with a matched period on the following day when the AFU was in operation. We were also able gain insights into the transport of aerosols around the ward by correlating the various PM signals from the respective sensors. We report our findings here to raise awareness about the ease with which aerosols can migrate in clinical settings, and also to demonstrate the potential benefits of employing supplementary room air filtration in ward spaces.

## Methods

### Ward layout and ventilation

The study took place in half of a ward, the layout of which and the locations of the AFU and sensors are shown in Figure 1. The ward, which was on the sixth floor, contained several bedded bays open to a central corridor and side rooms with doors to the corridor, which were frequently opened for patient care activities. The AFU was located in an open communal space within the ward. The study half of the ward comprised three side rooms and two bays, each with six beds. Sensor *A* was situated close to the AFU at a height of 2m (remaining sensors between 1.5m and 1.7m high; dictated by available electrical outlets). PM, CO_2_, temperature and RH data was collected automatically by the seven sensors which were wall mounted spaced around the ward as described in Figure 1 and Table 1.

**Figure 1.**
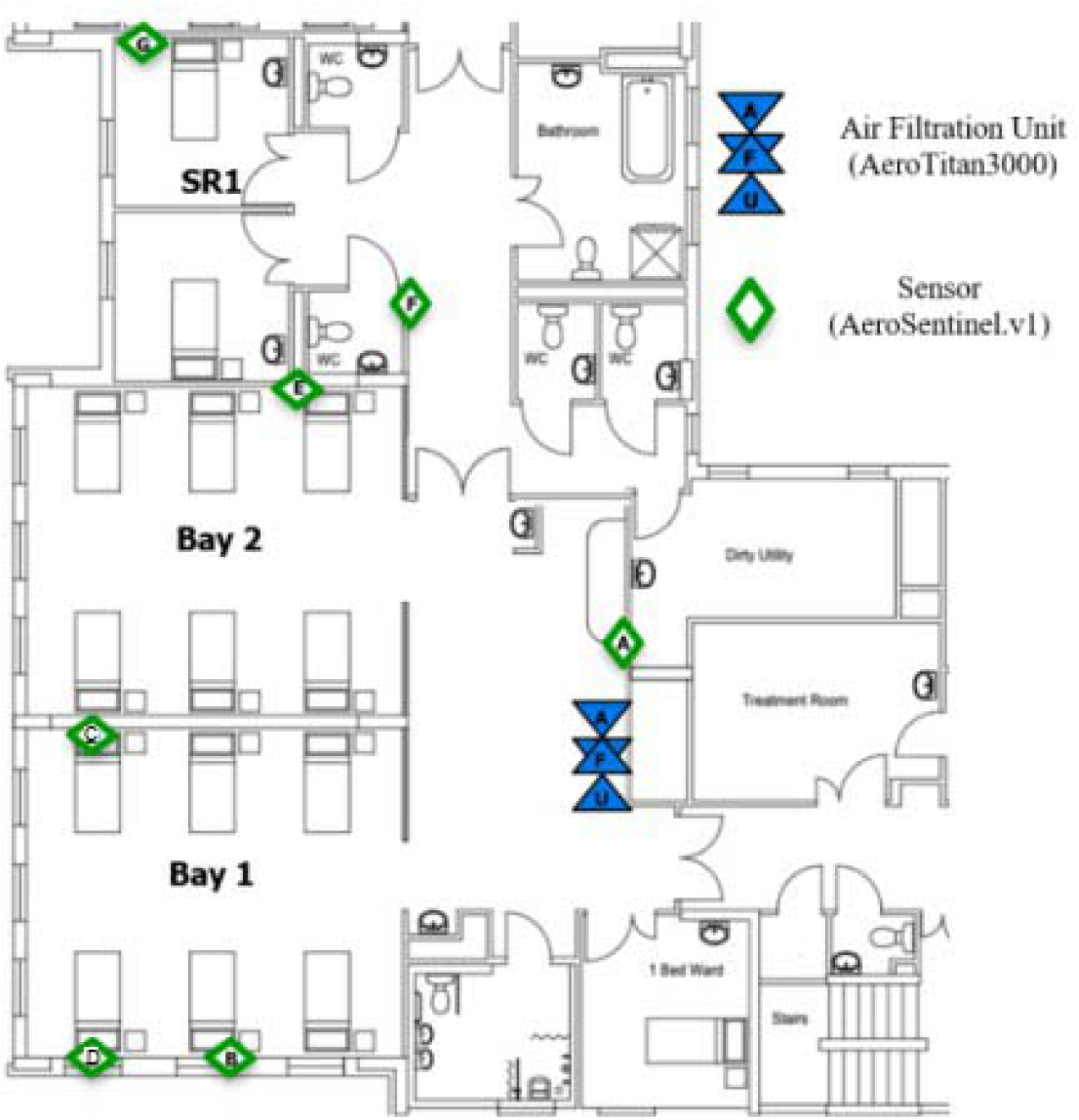
Layout of the medicine for older people ward showing the positions of the AFU and sensors.

**Table 1.** Locations of sensors on the ward.

| Sensor | Location |
| --- | --- |
| *A* | Communal space outside bays 1 & 2 |
| *B* | Bay 1 (Bed 2) |
| *C* | Bay 1 (Bed 3) |
| *D* | Bay 1 (Bed 4) |
| *E* | Bay 2 (Bed 6) |
| *F* | Corridor between bay 2 & side room E |
| *G* | Side Room 1 (SR1) |

The ward was ventilated by the combination of a central ducted mechanical ventilation system and openable aluminium sash windows, with the bed bays and side rooms being positively pressurised with respect to the central corridor. Historical records (measured by the hospital estates department) suggested that the ward ventilation rates ranged from 1.7 - 5.8 air changes per hour (ACH), with the median being 4.0 ACH.

### Ward operation

For both days included in this natural experiment (3^rd^ and 4^th^ August) the daytime nursing shift was from 07:30 to 19:30, with breakfast, lunch and dinner commencing at approximately 08:00, 12:00 and 17:00 respectively. The daily multidisciplinary team meeting commenced at approximately 08:45 on each day, in a room outside the study area, and ward rounds then commenced at approximately 09:15-09:30. The communal space where the AFU was situated was utilised daily by staff who congregated there to plan care activities as part of the ward round (Figure 2). Consequently, this was usually the most densely populated area of the ward.

**Figure 2.**
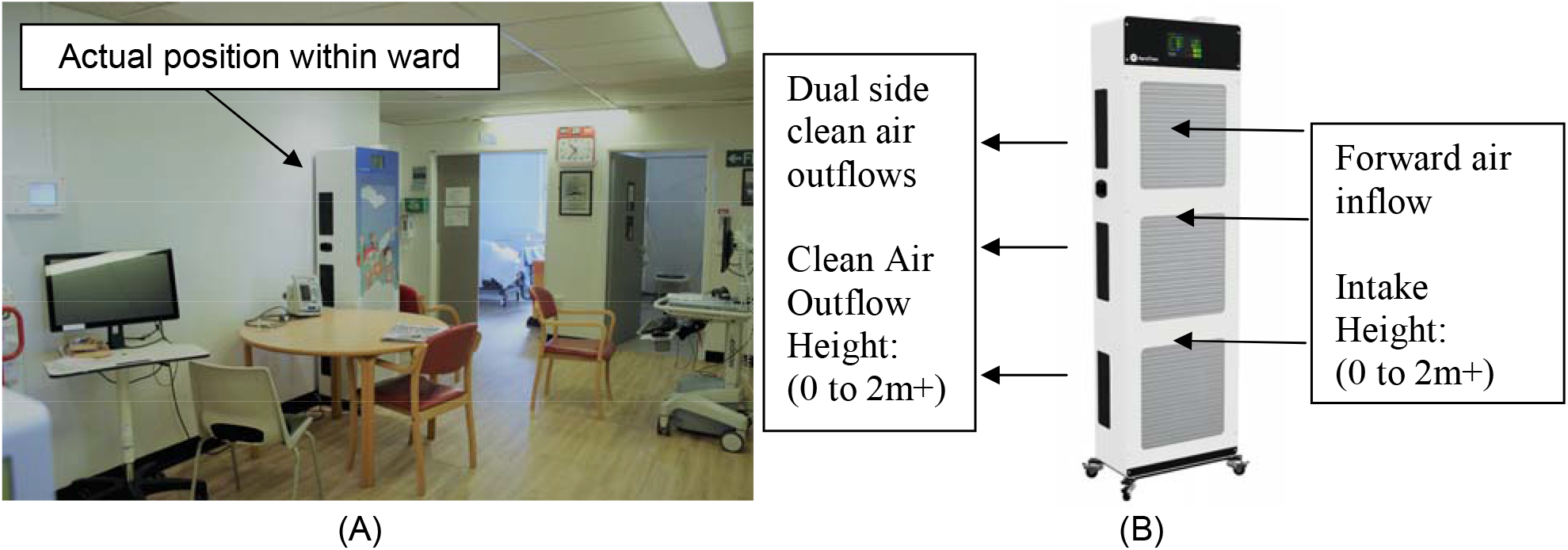
(A) Photograph showing the location of the AFU in the ward, and (B) detail showing the air intakes and outlets of the AFU.

With regard to the operation of the AFU, this was accidentally switched off early in the morning of the 3^rd^ August – a mistake that was rectified in the afternoon of the same day, some time after 15:00 when the device was restarted. Immediately prior to being switched off, the ADU had been operating on speed setting 2, which is its night-time default setting. On being re-started the ADU was inadvertently operating at speed setting 2, a mistake that was corrected on the morning of 4^th^ August when it was adjusted to speed setting 3.

### Air filtration unit (AFU)

A single AFU (AeroTitan3000; AirPurity UK Ltd, Cambridge UK) was sited opposite the two six bedded patient bays (Figures 1 and 2). To maximise the impact of the AFU, care was taken to ensure that the clean air discharge velocity was high to promote good air mixing throughout the ward space. Performance details of the AFU are presented in Table 2 including fan delivery rates for the various speed settings. The AFU was a hybrid encapsulated system which combined a series of HEPA filters and UV-C lamps (at 254 nm) over multiple heights and large surface areas to control the environment. The AFU was designed to manipulate the indoor environment by creating air currents resulting in both proximal and distant (>10m) cleaning and dilution through high laminar flow.

**Table 2.**
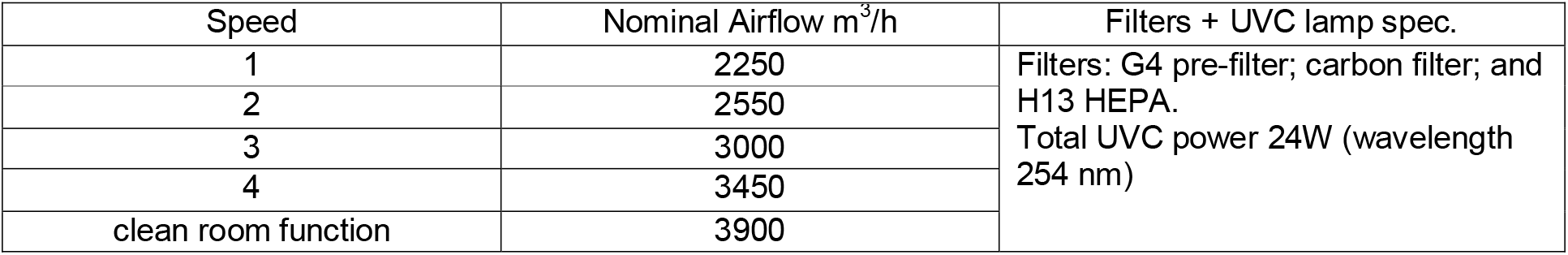
AFU Outline Specification

| Speed | Nominal Airflow m<sup>3</sup>/h | Filters + UVC lamp spec. |
| --- | --- | --- |
| 1 | 2250 | Filters: G4 pre-filter; carbon filter; and H13 HEPA.<br>Total UVC power 24W (wavelength 254 nm) |
| 2 | 2550 |  |
| 3 | 3000 |  |
| 4 | 3450 |  |
| clean room function | 3900 |  |

### Sensors

A group of sensors (AeroSentinel.v1; AirPurity UK Ltd, Cambridge UK) were used to relay environmental data back into a remote cloud system at 5 second intervals. These recorded the environmental data with the following accuracies: PM1 and PM2.5 (±10 μg/m^3^); PM4 and PM10 (±25 μg/m^3^); CO_2_ (±30 ppm); temperature (±0.4°C); and RH (±3%).

### Data

The data collected comprised time-series signals from the seven automatic sensors located on the ward as shown in Figure 1 and described in Table 1. The collected data from the various sensors was sampled every 1 minute, giving a total of 2782 data points per sensor over the two-day period. Across all the sensors a total of 7.6% of the data was missing. This missing data was imputed as being the mean value of the adjacent data points. For each sensor the data extracted is as shown in Table 3, with the PM fractions (i.e. the PM mass between the various intervals), hereafter simply referred to as PM1, PM2.5, PM4 and PM10 respectively.

**Table 3.** Signals extracted from the sensors on the ward.

| Signal | Description | Abbreviation | Units |
| --- | --- | --- | --- |
| PM1 fraction | Particle diameter < 1 µm | PM1 | µg/m<sup>3</sup> |
| PM2.5 fraction | Particle diameter between 1 and 2.5 µm | PM2.5 | µg/m<sup>3</sup> |
| PM4 fraction | Particle diameter between 2.5 and 4 µm | PM4 | µg/m<sup>3</sup> |
| PM10 fraction | Particle diameter between 4 and 10 µm | PM10 | µg/m<sup>3</sup> |
| CO<sub>2</sub> | Carbon dioxide level | CO<sub>2</sub> | ppm |
| Temperature | Air temperature | T | °C |
| Humidity | Relative humidity | RH | % |

### Vapour pressure

As well as monitoring PM1, PM2.5 and CO_2_, the sensors also recorded air temperature and RH. However, as RH is a relative value which is a function of temperature and vapour pressure (VP), in order to perform meaningful analyses it was necessary to compute the VP signal from the temperature and RH signals. This was done using the equations (1) and (2) below.

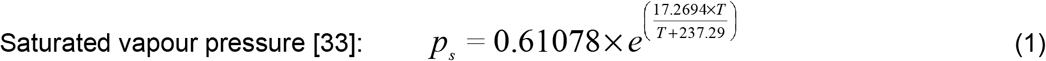

Where; *p*_*s*_ is saturated vapour pressure (kPa) and *T* is air temperature (°C).

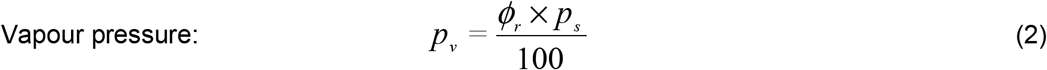

Where; *p*_*v*_ is vapour pressure (kPa) and *ϕ*_*r*_ is relative humidity (%).

### Change point analysis and validation

As the work reported here is an after-the-fact study of a natural experiment that occurred due to unforeseen circumstances, care was taken to minimise the use of any *a priori* assumptions. For example, because contemporaneous records were not kept, no assumptions were made regarding the precise points in time when the AFU was switched off and on, or when the speed settings were changed. These were not recorded at the time and could only be confirmed after-the-fact by extracting the memory cards from the AFU, something that we reserved for validation purposes only. Consequently, when analysing the data we decided to employ a change point methodology to identify natural breaks in the time-series data – a process which required no *a priori* information.

Change point analysis is a technique which seeks to identify points in time where structural changes have occurred in a time-series signal [34, 35]. As such, it is frequently used to retrospectively identify when events have occurred. One commonly used approach in change point analysis is to identify when the mean of the signal has significantly altered [36]. Accordingly, we used the ‘cpt.mean’ function in the ‘changepoint’ package in R, which utilised a pruned exact linear time (PELT) methodology [36] to identify break points where the mean of the PM1 signal from sensor *A* structurally changed. This sensor which was selected because it was located in the communal space, near to the AFU and as such was ideally placed to monitor both changes in occupancy level as well as the operation of the AFU.

The aim of the change point analysis was to identify (from the PM1 signal alone) points in time where structural changes occurred, so that these could be compared with events that happened on the ward. For simplicity, minor change points that occurred when the PM1 signal was transient during the period in which the AFU was not in operation were ignored. This was because the PM levels were not controlled when the AFU was off, resulting in large fluctuations in the PM1 signal and considerable transience, which led to multiple mini-change points being identified during this period, as illustrated in Figure 3. Change points were therefore deemed to have occurred at all points where major changes in the structural mean of the signal happened. These were identified by visual inspection and statistically tested by applying the Chow test, with p<0.05 deemed to be significant.

**Figure 3.**
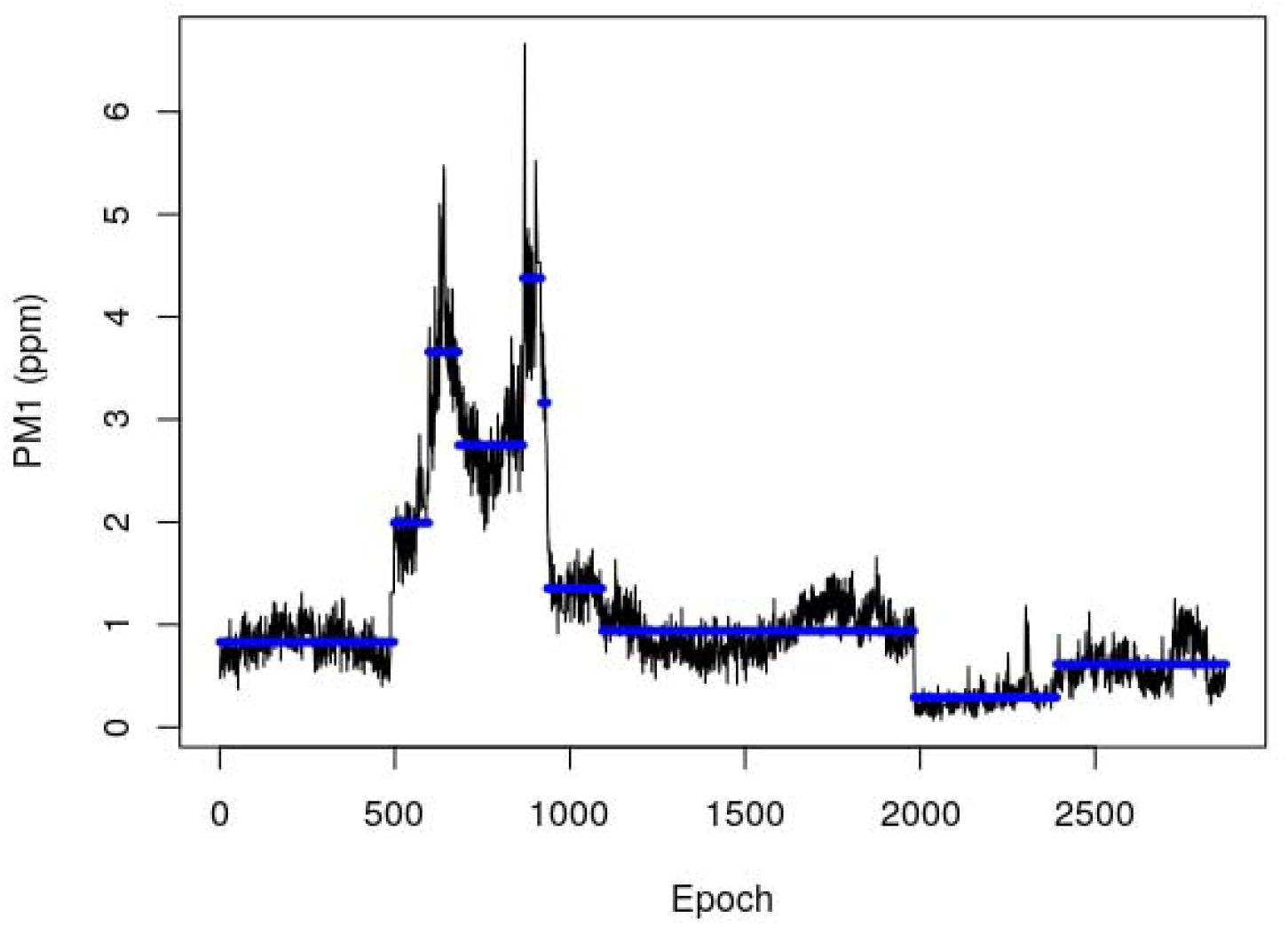
Results of change point analysis showing the PM1 signal from sensor *A* (black), with the interrupted mean signal (blue) for each section. Change points were identified at epochs 497, 596, 681, 866, 918, 935, 1092, 1982 and 2389.

### Primary statistical analysis

The hypothesis tested was that the respective signal levels were higher when the AFU was not in operation on the 3^rd^ August compared with a matched period on the 4^th^ August when the AFU was in operation. This was tested using an unpaired Mann-Whitney test in R (R Core Team (2021). URL https://www.R-project.org/). The observed effect size was evaluated using Cliff’s delta statistic, with the magnitude assessed using the thresholds provided by Romano et al [37] (i.e. |δ| < 0.147 deemed negligible; 0.147 ≤ |δ| < 0.33 deemed small; 0.33 ≤ |δ| < 0.474 deemed medium, and 0.474 ≤ |δ| deemed large).

To assess the relationships between the signals from the individual sensors, the Pearson correlation r values were computed together with their statistical significance. This was done for each individual sensor using the entire data set over the entire two-day study period.

In addition, the extent to which airborne PM might be migrating around the ward was assessed by analysing the between sensor correlation using only the data collected when the AFU was not in operation on the 3^rd^ August.

For all tests, p<0.05 was deemed as being significant.

### Secondary post hoc analysis

Having conducted the initial change-point analysis and tested the primary hypothesis, the results suggested that further secondary *post hoc* analysis was worth pursuing. This involved *post hoc* testing of a secondary hypothesis, namely that increasing the AFU speed setting from 2 to 3 on the 4^th^ August reduced the levels of the respective signals from the various sensors. This was tested using an unpaired Mann-Whitney test, with p<0.05 deemed as being significant. The effect size was assessed using Cliff’s delta statistic.

In order to estimate the additional air change rate provided by the AFU, post hoc decay analysis was performed on the PM1 signal collected from sensor *A* near to the AFU. This involved analysing a section of the signal from epoch 880 (14:40 on 3^rd^ August) to epoch 1000 (16:40 on 3^rd^ August), which included the point in time when the AFU was switched on again. The decay analysis was performed using equation 3 below.

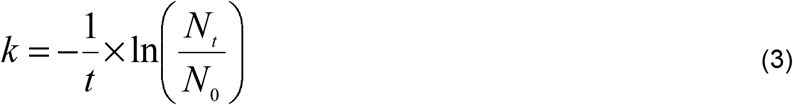

Where: *N*_*t*_ is the number of aerosol particles in the air at time *t* hours; *N*_*0*_ is the number of aerosol particles in the air at time *t* = 0 hours (i.e. the initial state); *t* is time in hours; and *k* is the decay constant representing the rate at which particles are removed from the air (h^-1^). The decay rate constant, *k*, is generally expressed in terms of equivalent air changes per hour (AC/h) for air cleaning devices.

Using Equation 3, it is possible to determine the particle removal rate experimentally by simply measuring the time taken for the particle concentration in the air to fall by a certain pre-determined fraction. This fraction can be any value, but by convention is usually the time taken for the particle concentration in the air to reduce by half.

## Results

### Change point analysis results

Five major change points were identified across the time period examined (Table 4, figures 3 and 4). Of these, CP1, CP2 and CP4 closely coincided with the time-stamps that were recovered from the AFU memory card in December 2021 (Table 4), namely: 08:16 (epoch 496) for the device being turned off; 15:34 (epoch 934) for the device being restarted on speed setting 2; and 09:05 (epoch 1985) for the speed setting being increased from 2 to 3. However, due to technical difficulties (missing data points, etc.), the recovered time-stamp data had to be estimated and therefore can only be considered approximate only, and indicative of points in time where changes occurred. Details of the computed change points and the respective validation time-stamps are shown in Table 4.

**Table 4.**
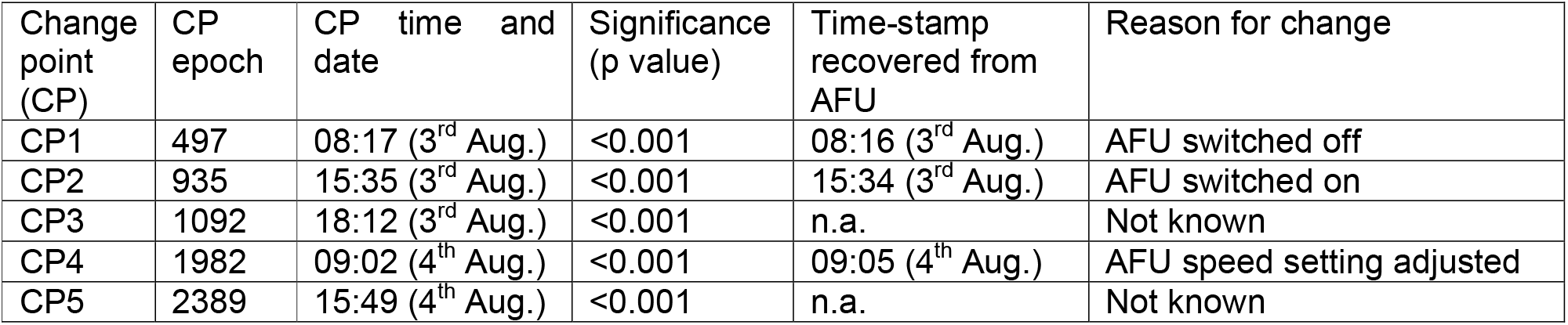
Occurrence of major change points in PM1 signal for sensor *A*, together with time-stamp data recovered from the air disinfection unit (AFU).

**Figure 4.**
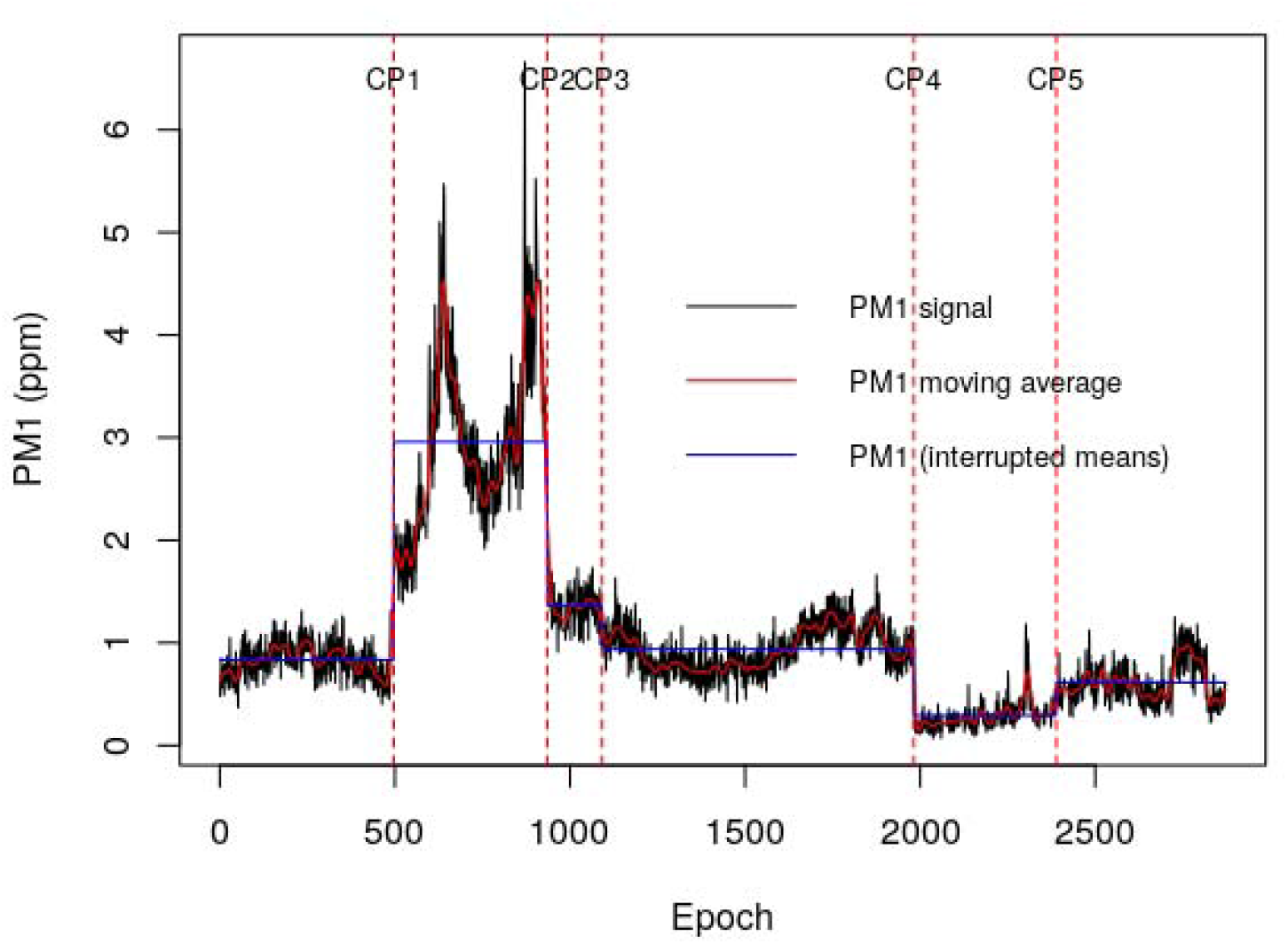
PM1 signal from sensor *A* (black) with the interrupted mean signal (blue) for the sections between the major change points. Major change points were identified at epochs 497, 935, 1092, 1982 and 2389.

### Time series results

The collated time-series results are presented in figures 5-9, which show the respective PM and CO_2_ signals from all the sensors for 3^rd^ and 4^th^ August, together with the change points and meal times mapped on top. Examining the particulates across the range of sizes measured reveals similar patterns to the PM1 signal shown in figures 3 and 4, whilst comparison of PM data from all sensors demonstrated large increases in particulates throughout the ward when the AFU was not operational. This widespread pattern of particulate counts implies both the free movement of aerosols between sub-compartments (ward bays etc.) and an impact of air filtration beyond the immediate environment of the AFU itself (>10m). Although the AFU had an impact on all particulate sizes, its effect was particularly marked in the larger sizes (PM4 and PM10), which were rendered almost undetectable during AFU operation. Notably, CO_2_ levels showed a similar change (Figure 9), implying the AFU possibly had an impact on overall ward ventilation. Overlaying the times for breakfast, lunch and dinner illustrates the diurnal variation in particulates and CO_2_ and suggests that human activity is the major driver of aerosol generation on the ward. This is evident from the signal spikes that occurred during the daytime on both 3^rd^ and 4^th^ August. Having said this, it is noticeable that for all particulate sizes the variance in the PM signals greatly reduced when the AFU was in operation.

**Figure 5.**
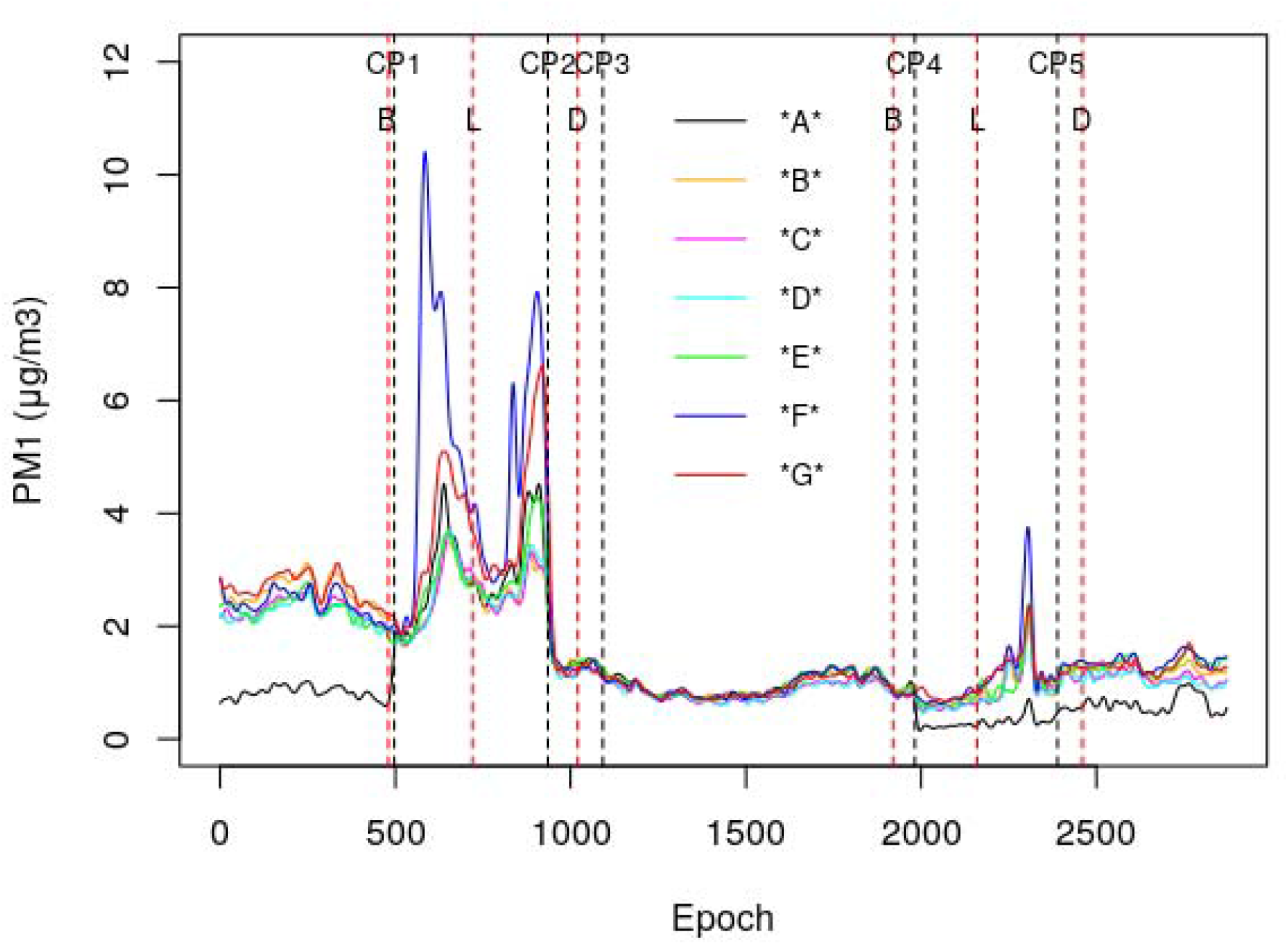
Collated smoothed PM1 signals from all the sensors for 3^rd^ and 4^th^ August. The black dashed lines denote where the change points (CPs) occurred and the dashed red lines denote when breakfast (B), lunch (L) and dinner (D) were served to the patients on the ward. (Noise from the PM signals was removed using a cubic smoothing spline in R (R Core Team (2021). URL https://www.R-project.org/.), with the smoothing parameter set to 0.1.)

**Figure 6.**
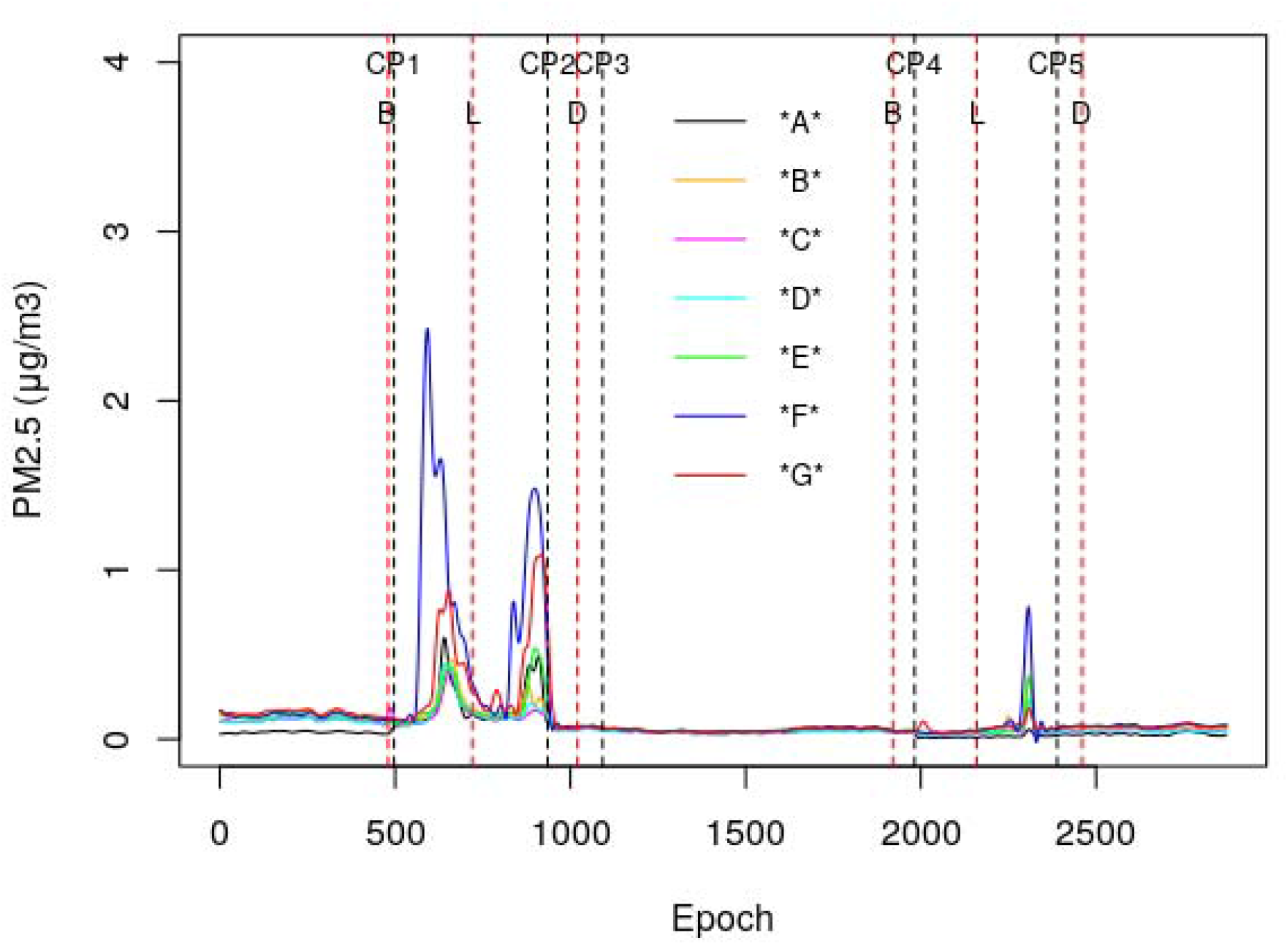
Collated smoothed PM2.5 signals from all the sensors for 3^rd^ and 4^th^ August. The black dashed lines denote where the change points (CPs) occurred and the dashed red lines denote when breakfast (B), lunch (L) and dinner (D) were served to the patients on the ward. (Noise from the PM signals was removed using a cubic smoothing spline in R (R Core Team (2021). URL https://www.R-project.org/.), with the smoothing parameter set to 0.1.)

**Figure 7.**
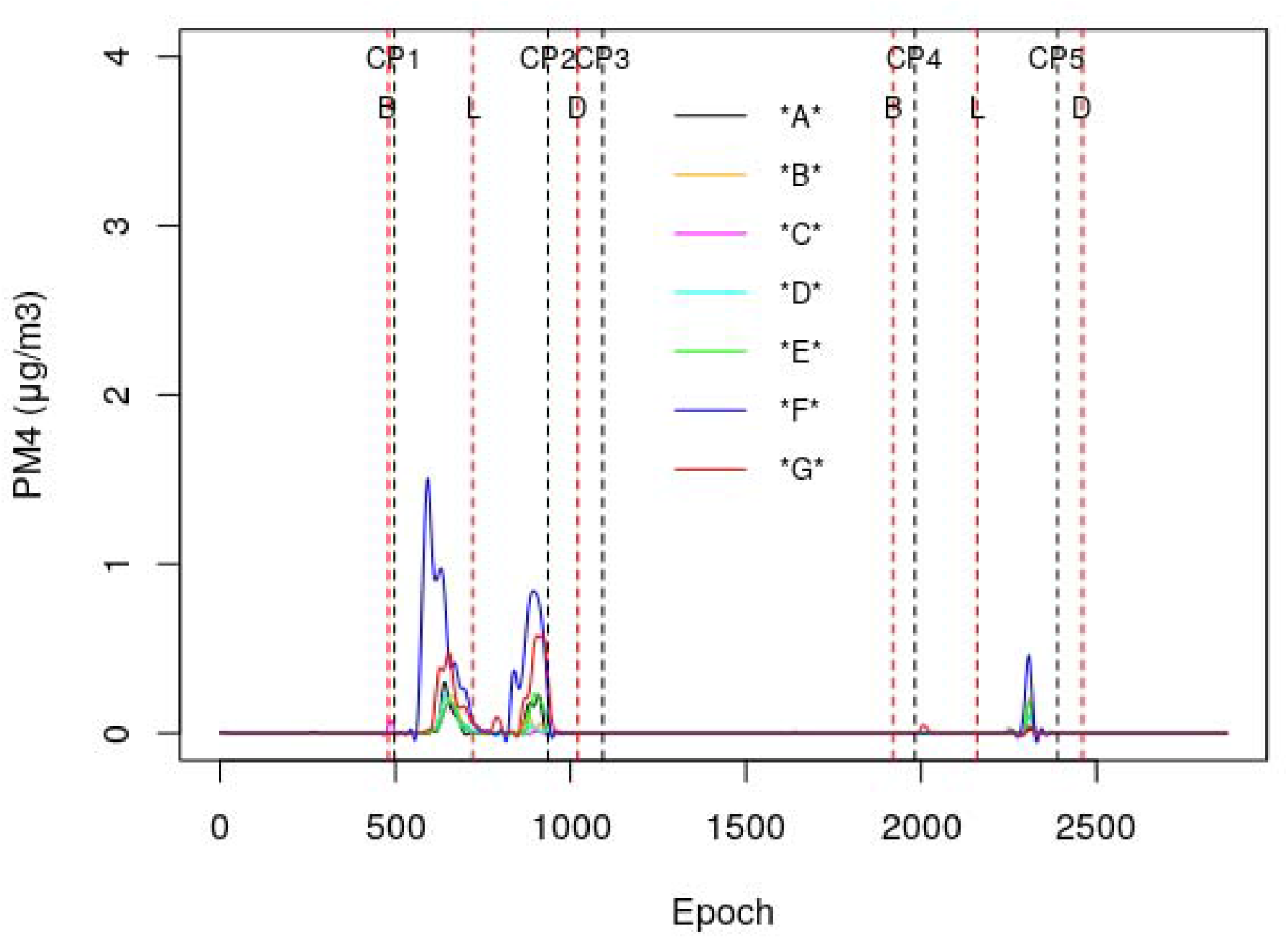
Collated smoothed PM4 signals from all the sensors for 3^rd^ and 4^th^ August. The black dashed lines denote where the change points (CPs) occurred and the dashed red lines denote when breakfast (B), lunch (L) and dinner (D) were served to the patients on the ward. (Noise from the PM signals was removed using a cubic smoothing spline in R (R Core Team (2021). URL https://www.R-project.org/.), with the smoothing parameter set to 0.1.)

**Figure 8.**
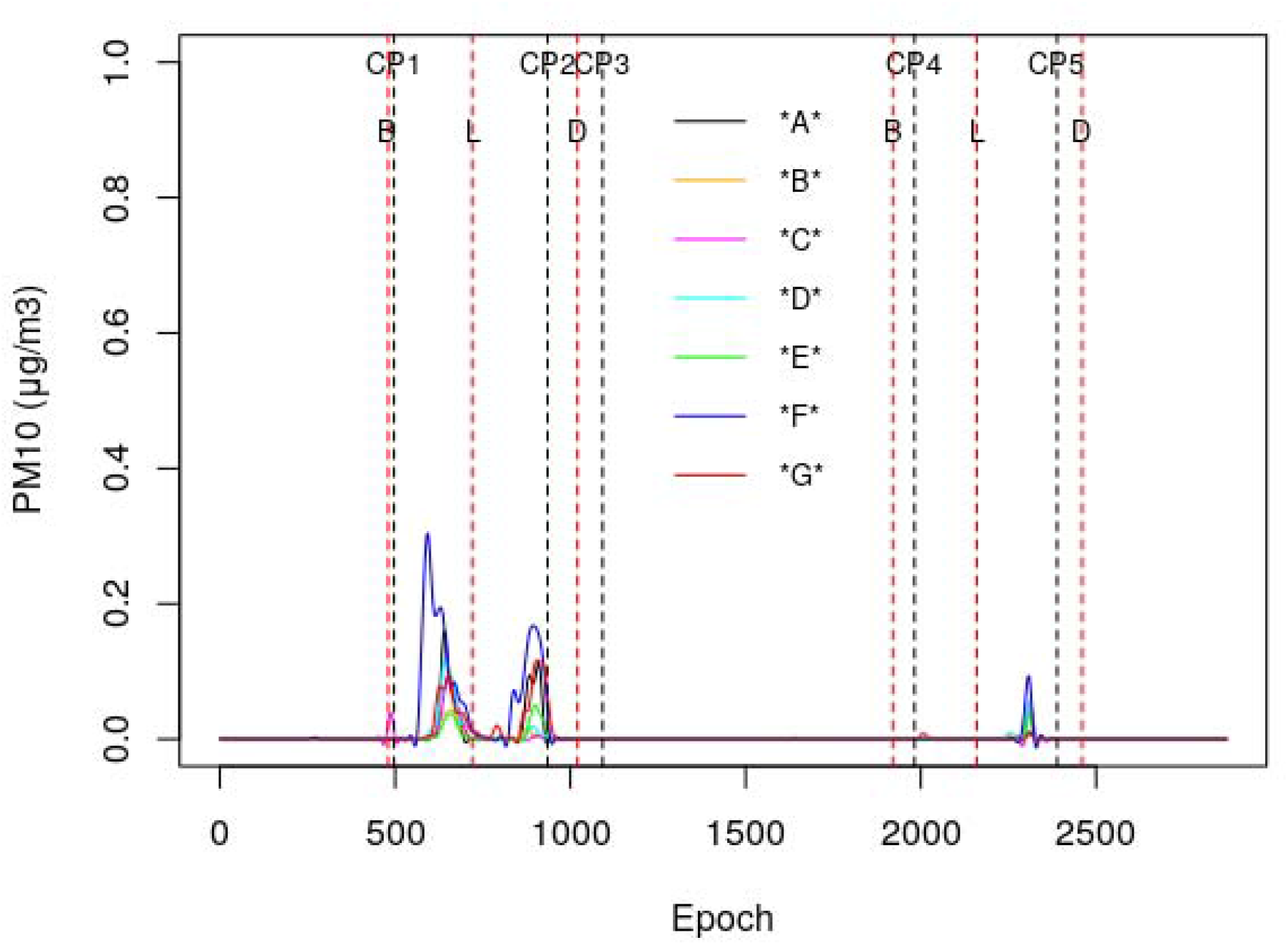
Collated smoothed PM10 signals from all the sensors for 3^rd^ and 4^th^ August. The black dashed lines denote where the change points (CPs) occurred and the dashed red lines denote when breakfast (B), lunch (L) and dinner (D) were served to the patients on the ward. (Noise from the PM signals was removed using a cubic smoothing spline in R (R Core Team (2021). URL https://www.R-project.org/.), with the smoothing parameter set to 0.1.)

**Figure 9.**
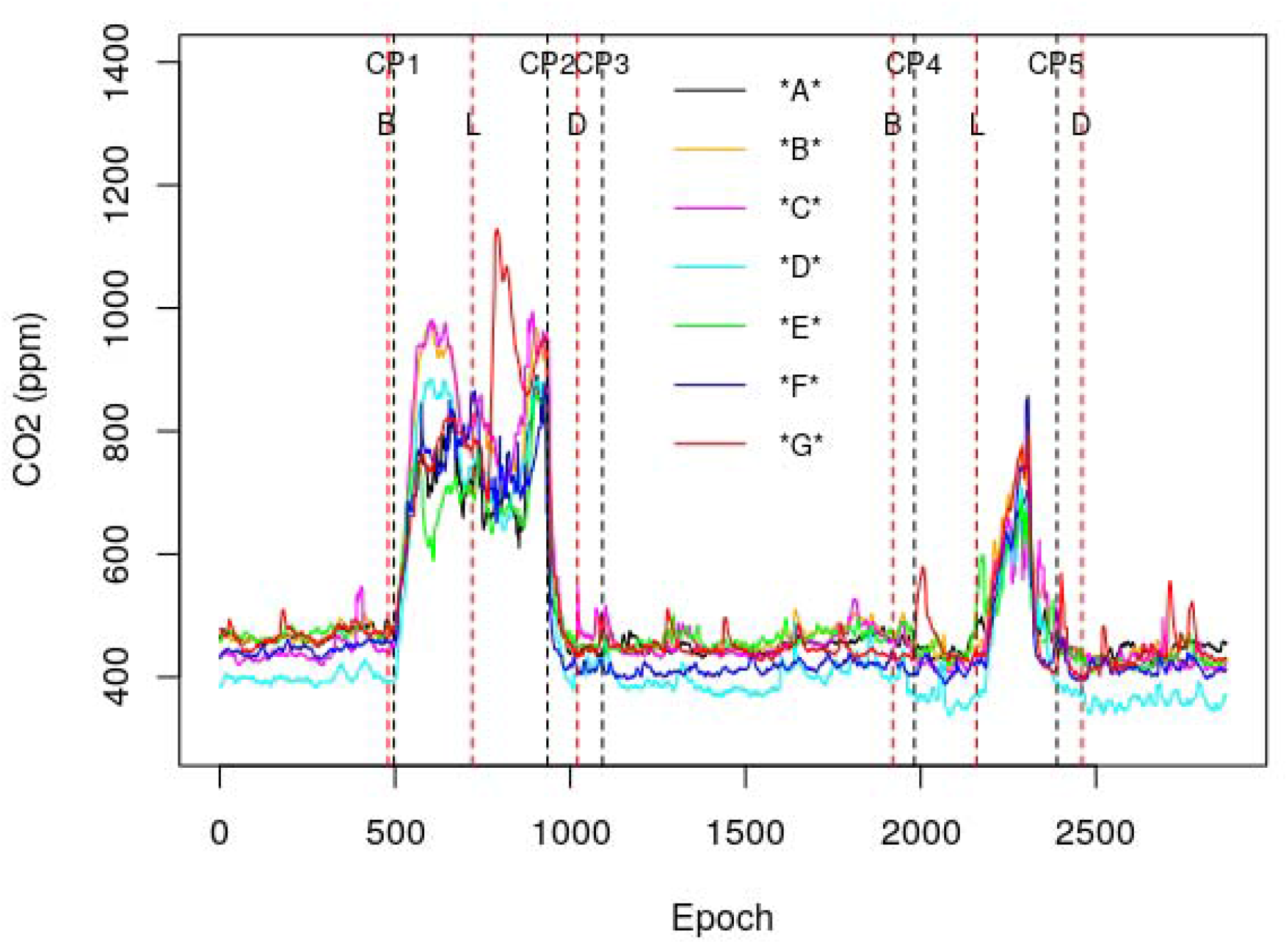
Collated CO_2_ signals from all the sensors for 3^rd^ and 4^th^ August. The black dashed lines denote where the change points (CPs) occurred and the dashed red lines denote when breakfast (B), lunch (L) and dinner (D) were served to the patients on the ward.

### Descriptive statistics results

The signals shown in figures 5-9 are summarized in Table S1 in the Supplementary Material, which shows the descriptive statistics for the signals from the various sensors. (NB. All sensors produced an identical vapour pressure signal). The results in Table S1 show the means and standard deviations for the different signals from the various sensors for the time periods between the identified change points. From this it can be seen that for all the sensors, the observed signal readings were much higher between CP1 and CP2, when the AFU was off, compared with the other periods when it was in operation.

### Statistical analysis results

The results of the statistical analysis to test the hypothesis that the AFU reduced the respective sensor signal levels are presented in Table 5. This hypothesis stated that the signal levels were higher when the AFU was not in operation (i.e. from CP1 to CP2 – 08:17 to 15:34 on the 3^rd^ August) compared with a matched period (08:17 to 15:34) on the 4^th^ August when the AFU was in operation.

**Table 5.** Results of statistical test of the hypothesis that the signal levels were higher when the AFU was not in operation on the 3^rd^ August compared with a matched period on the 4^th^ August when it was in operation. (NB. All results were strongly significant after Bonferroni correction.)

| Signal | Sensor ID | AFU off<br>Mean | AFU off<br>SD | AFU on<br>Mean | AFU on<br>SD | Significance<br>p-value | Effect size<br>Cliff's delta | Effect<br>magnitude |
| --- | --- | --- | --- | --- | --- | --- | --- | --- |
| PM1 | *G* | 3.728 | 1.299 | 1.026 | 0.418 | 6.83E-141 | 0.986 | large |
|  | *F* | 5.054 | 2.347 | 1.103 | 0.709 | 2.11E-134 | 0.963 | large |
|  | *E* | 2.827 | 0.696 | 0.893 | 0.399 | 6.72E-138 | 0.976 | large |
|  | *A* | 2.961 | 0.871 | 0.352 | 0.235 | 9.32E-145 | 1.000 | large |
|  | *B* | 2.607 | 0.572 | 0.896 | 0.434 | 6.91E-134 | 0.961 | large |
|  | *C* | 2.599 | 0.574 | 0.815 | 0.380 | 1.02E-138 | 0.979 | large |
|  | *D* | 2.641 | 0.613 | 0.820 | 0.385 | 1.58E-138 | 0.978 | large |
| PM2.5 | *G* | 0.394 | 0.308 | 0.063 | 0.032 | 1.86E-137 | 0.974 | large |
|  | *F* | 0.747 | 0.650 | 0.091 | 0.155 | 2.10E-123 | 0.922 | large |
|  | *E* | 0.217 | 0.125 | 0.061 | 0.066 | 8.15E-126 | 0.930 | large |
|  | *A* | 0.201 | 0.140 | 0.018 | 0.014 | 1.08E-145 | 0.999 | large |
|  | *B* | 0.198 | 0.106 | 0.064 | 0.071 | 1.31E-117 | 0.899 | large |
|  | *C* | 0.153 | 0.078 | 0.045 | 0.041 | 7.72E-126 | 0.930 | large |
|  | *D* | 0.168 | 0.094 | 0.047 | 0.041 | 1.06E-124 | 0.925 | large |
| PM4 | *G* | 0.146 | 0.194 | 0.004 | 0.012 | 3.31E-67 | 0.594 | large |
|  | *F* | 0.369 | 0.425 | 0.022 | 0.097 | 3.38E-72 | 0.627 | large |
|  | *E* | 0.044 | 0.073 | 0.008 | 0.038 | 9.93E-28 | 0.326 | small |
|  | *A* | 0.046 | 0.085 | 0.001 | 0.004 | 3.87E-23 | 0.266 | small |
|  | *B* | 0.039 | 0.066 | 0.010 | 0.043 | 1.89E-24 | 0.318 | small |
|  | *C* | 0.021 | 0.047 | 0.004 | 0.023 | 9.74E-18 | 0.224 | small |
|  | *D* | 0.031 | 0.058 | 0.005 | 0.020 | 4.27E-27 | 0.342 | medium |
| PM10 | *G* | 0.030 | 0.039 | 0.001 | 0.003 | 2.42E-64 | 0.550 | large |
|  | *F* | 0.074 | 0.086 | 0.004 | 0.019 | 6.57E-71 | 0.600 | large |
|  | *E* | 0.009 | 0.015 | 0.002 | 0.008 | 1.29E-22 | 0.263 | small |
|  | *A* | 0.024 | 0.044 | 0.001 | 0.003 | 4.63E-24 | 0.263 | small |
|  | *B* | 0.007 | 0.013 | 0.002 | 0.008 | 2.66E-18 | 0.237 | small |
|  | *C* | 0.011 | 0.025 | 0.002 | 0.012 | 2.93E-16 | 0.204 | small |
|  | *D* | 0.017 | 0.031 | 0.003 | 0.011 | 5.27E-26 | 0.326 | small |
| CO2 | *G* | 815.175 | 139.885 | 513.280 | 109.663 | 4.96E-118 | 0.902 | large |
|  | *F* | 734.244 | 80.023 | 479.591 | 111.878 | 1.49E-112 | 0.880 | large |
|  | *E* | 691.243 | 72.783 | 501.255 | 72.280 | 6.77E-123 | 0.920 | large |
|  | *A* | 713.817 | 80.280 | 499.449 | 78.060 | 1.23E-120 | 0.912 | large |
|  | *B* | 818.856 | 115.035 | 520.789 | 105.350 | 1.41E-124 | 0.927 | large |
|  | *C* | 833.165 | 121.853 | 494.870 | 85.350 | 2.84E-130 | 0.948 | large |
|  | *D* | 748.612 | 113.939 | 447.098 | 102.953 | 1.09E-124 | 0.927 | large |
| VP | All sensors | 1.368 | 0.051 | 1.179 | 0.063 | 4.90E-139 | 0.980 | large |

From Table 5 it can be seen that for all the sensors the action of the AFU was associated with a large effect on PM1, PM2.5, CO_2_ and VP levels, which was highly significant. This strongly suggests that for the respective matched periods on the 3^rd^ and 4^th^ August, when the ward was busy, the action of the AFU was associated with large reductions in the level of these signals. By comparison, although statistically significant reductions were also observed for the PM4 and PM10 signals, the effect size for many of the sensors was much smaller, reflecting the lower particle counts in this size range and the fact that these large particles tend to settle out of the air at a faster rate. As such, the hypothesis can be accepted for all the signals.

When Pearson correlation analysis was performed on data collected from the individual sensors, high positive correlations were observed between most of the signals. This is well illustrated in Figure 10 and Table 6, which show the respective signals and correlations for sensor *G*. From these it can be seen that all the PM signals are strongly correlated with each other (i.e. r>0.7). The CO_2_ and VP are also positively correlated with the PM signals and each other, although not as strongly.

**Table 6.** Pearson correlation r values for signals from sensor *G* over the entire two-day study period.

|  | PM1 | PM2.5 | PM4 | PM10 | CO2 | VP |
| --- | --- | --- | --- | --- | --- | --- |
| PM1 | 1.000 | 0.884 | 0.718 | 0.719 | 0.713 | 0.732 |
| PM2.5 | 0.884 | 1.000 | 0.959 | 0.958 | 0.675 | 0.554 |
| PM4 | 0.718 | 0.959 | 1.000 | 0.996 | 0.578 | 0.387 |
| PM10 | 0.719 | 0.958 | 0.996 | 1.000 | 0.582 | 0.390 |
| CO2 | 0.713 | 0.675 | 0.578 | 0.582 | 1.000 | 0.559 |
| VP | 0.732 | 0.554 | 0.387 | 0.390 | 0.559 | 1.000 |

NB. All correlations significant at $p < 0.001$ .

**Figure 10.**
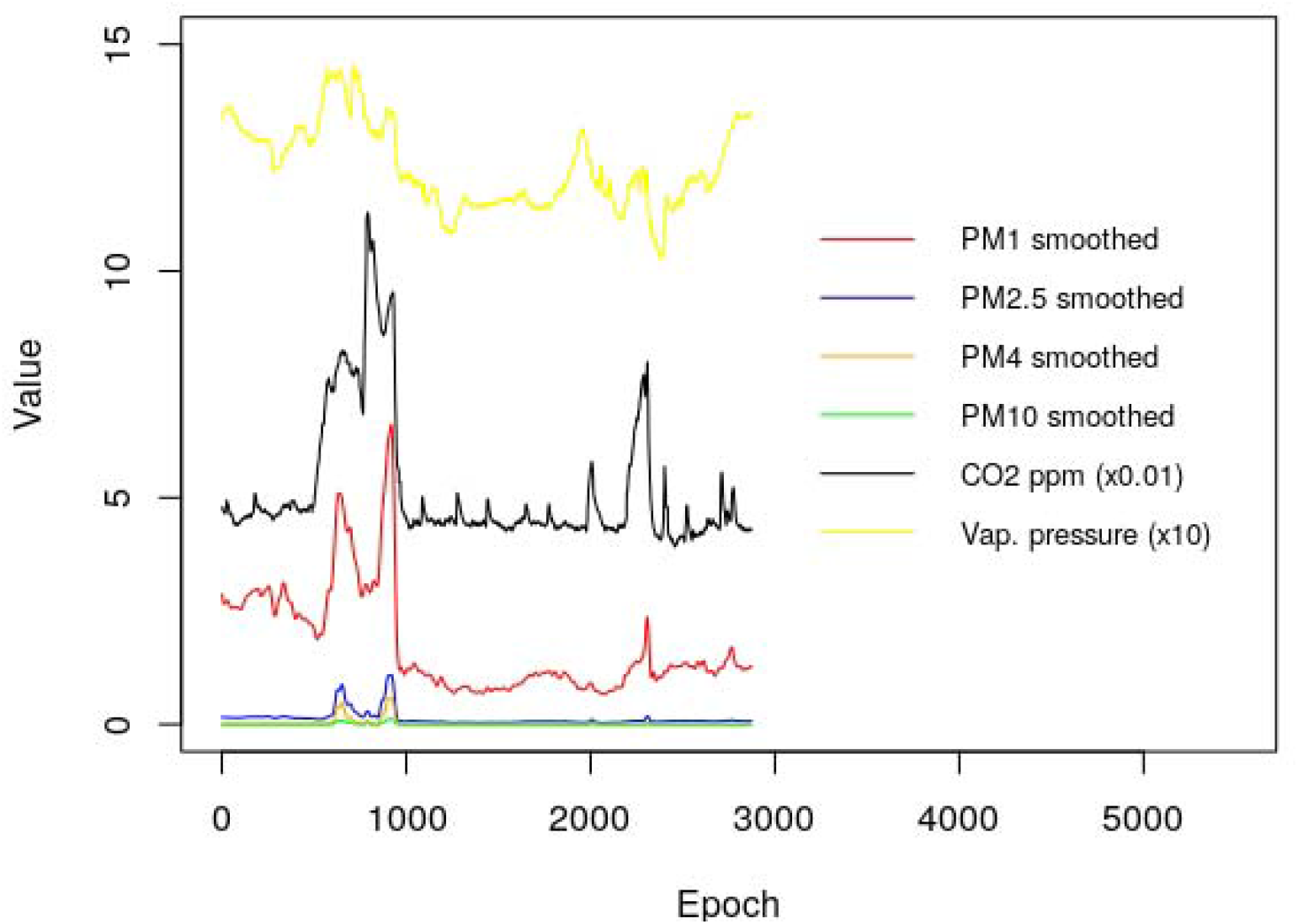
Smoothed PM, CO_2_ and VP signals from sensor *G* in the ward side room.

Correlation analysis of the PM, CO_2_ and VP signals from the other sensors revealed similar r value ranges as follows: sensor *A* (r = 0.315 – 0.998; all p<0.001); sensor *B* (r = 0.292 – 0.973; all p<0.001); sensor *C* (r = 0.232 – 0.996; all p<0.001); sensor *D* (r = 0.337 – 0.992; all p<0.001); sensor *E* (r = 0.279 – 0.986; all p<0.001); and sensor *F* (r = 0.448 – 0.999; all p<0.001). More specifically, for the relationship between CO_2_ and the various PM signals, the correlation means and ranges were: PM1 (r = 0.690 (0.551 – 0.817); all p<0.001); PM2.5 (r = 0.682 (0.604 – 0.755); all p<0.001); PM4 (r = 0.531 (0.347 – 0.639); all p<0.001); and PM10 (r = 0.525 (0.343 – 0.636); all p<0.001). Collectively, this indicates that over the whole study period the PM and CO_2_ signals from the individual sensors in the ward space tended to rise and fall together, with CO_2_ most strongly correlated with the smaller aerosols.

One surprising finding is that the action of the AFU greatly reduced CO_2_ levels throughout the ward when in operation. From Figure 9 it can be seen that this effect was not localised, but occurred simultaneously in all the sub-compartments of the ward. This effect is difficult to explain, but may be due to a combination of better mixing of the ward air and increased entrainment of fresh air from outside through the windows due to the higher room air velocities when the AFU was in operation.

Another unexpected finding was that for many of the sensors, the ward vapour pressure (VP) was positively correlated with both the PM signals and CO_2_, as can be seen in Table 6, which shows the correlations between the signals from sensor *G*, located in a side room with a door that could be closed. From this it can be seen that for this sensor, VP was most strongly correlated with PM1 (r = 0.732, p<0.001), compared with PM2.5 (r = 0.554, p<0.001) and CO_2_ (r = 0.559, p<0.001). However, other sensors exhibited much weaker VP correlations, with for example sensor *B* (located in Bay 1) the correlation involving VP ranging from r = 0.123 for PM1 (p = 0.01) to r = 0.332 for PM10 (p<0.001). Indeed, for sensor *E* none of the correlations between VP and the PM signals reached significance.

The between-sensor correlations for the matching PM and CO_2_ signals from the respective sensors when the AFU was not in operation were also very strong. This is illustrated in Table 7, which shows the correlations between the PM1 signals from the respective sensors, all of which were highly significant (p<0.001). From this it can be seen that the correlations between all the sensor PM1 signals were all very strong, with the possible exception of those for sensor *F* located in the corridor, which tended to exhibit weaker correlations.

**Table 7.** Pearson correlation r values for the PM1 signals from respective sensors over the period for which the AFU was not in operation.

|  | *G* | *F* | *E* | *A* | *B* | *C* | *D* |
| --- | --- | --- | --- | --- | --- | --- | --- |
| *G* | 1 | 0.540 | 0.868 | 0.834 | 0.764 | 0.753 | 0.771 |
| *F* | 0.540 | 1 | 0.552 | 0.54 | 0.386 | 0.343 | 0.371 |
| *E* | 0.868 | 0.552 | 1 | 0.816 | 0.758 | 0.765 | 0.772 |
| *A* | 0.834 | 0.54 | 0.816 | 1 | 0.791 | 0.723 | 0.759 |
| *B* | 0.764 | 0.386 | 0.758 | 0.791 | 1 | 0.838 | 0.822 |
| *C* | 0.753 | 0.343 | 0.765 | 0.723 | 0.838 | 1 | 0.844 |
| *D* | 0.771 | 0.371 | 0.772 | 0.759 | 0.822 | 0.844 | 1 |

NB. All correlations significant at $p < 0.001$ .

Between-sensor analysis of the other signals for the period when the AFU was not in operation (i.e. between CP1 and CP2) revealed strong positive correlations between the PM signals from all the sensors, except for sensor *F* which exhibited noticeably weaker correlations. Excluding sensor *F*, the r value ranges between the various sensors were: PM2.5 (r = 0.560 – 0.904; all p<0.001); PM4 (r = 0.434 – 0.891; all p<0.001); and PM10 (r = 0.442 – 0.886; all p<0.001). As such, this indicates that the signals from most of the sensors in the ward space tended to rise and fall together, suggesting that when the AFU was not in operation, aerosols of various sizes were freely migrating between the various zones within the ward. The equivalent r value ranges for sensor *F* were somewhat lower (PM2.5 (r = 0.239 – 0.473; all p<0.001); PM4 (r = 0.189 – 0.425; all p<0.001); and PM10 (r = 0.184 – 0.421; all p<0.001), although still strongly significant.

Analysis of the relationship between CO_2_ and the various PM signals during the period when the AFU was not in operation revealed the correlation means and ranges to be: PM1 (r = 0.514 (0.394 – 0.624); all p<0.001); PM2.5 (r = 0.463 (0.300 – 0.616); all p<0.001); PM4 (r = 0.410 (0.263 – 0.577); all p<0.001); and PM10 (r = 0.401 (0.264 – 0.570); all p<0.001). Although these correlations are weaker than those exhibited for the whole study period, they still indicate that a close relationship existed between CO_2_ and the various PM signals, particularly the smaller aerosols, during the period when the AFU was off.

### Post hoc analysis results

The results of the *post hoc* analysis to test whether or not increasing the AFU speed setting from 2 (i.e. from CP2 to CP4) to 3 (i.e. after CP4) had any effect on the PM, CO_2_ and VP signal levels are presented in Table 8. From this it can be seen that a complex picture emerges. While increasing the fan speed of the AFU from setting 2 to 3 clearly resulted in a large significant reduction in the PM1 and PM2.5 counts recorded by sensor *A* (i.e. the sensor located closest to the AFU), the corresponding counts for the other sensors all significantly increased (although the magnitude of this effect was negligible), as did the PM4 and PM10 counts for all the sensors. This indicates that apart from cleaning the air in the vicinity of the AFU, increasing the fan speed from 2 to 3 had little impact on PM levels elsewhere on the ward. Indeed, in most locations the PM counts actually marginally increased when the speed setting was changed at 9:02 on the 4^th^ August, something that probably reflected increased ward activity during the daytime compared with night-time. Interestingly, increasing the speed setting did lead to a modest reduction in CO_2_ levels for some of the sensors, but noticeably not for sensor *A*.

**Table 8.** Results of the *post hoc* statistical test to determine whether or not the signal levels were lowered when the AFU speed setting was increased from 2 to 3 at CP4 on the 4^th^ August. (NB. All results were strongly significant after Bonferroni correction, except those marked * which did not reach significance.)

| Signal | Sensor ID | Speed 2<br>Mean | Speed 2<br>SD | Speed 3<br>Mean | Speed 3<br>SD | Significance<br>p-value | Effect size<br>Cliff's delta | Effect<br>magnitude |
| --- | --- | --- | --- | --- | --- | --- | --- | --- |
| PM1 | *G* | 0.984 | 0.374 | 1.179 | 0.339 | 1.30E-56 | -0.417 | medium |
|  | *F* | 0.999 | 0.298 | 1.262 | 0.527 | 7.34E-50 | -0.391 | medium |
|  | *E* | 1.010 | 0.274 | 1.126 | 0.379 | 2.76E-14 | -0.200 | small |
|  | *A* | 1.002 | 0.266 | 0.466 | 0.236 | 7.79E-237 | 0.865 | large |
|  | *B* | 1.014 | 0.268 | 1.073 | 0.366 | 2.70E-05 | -0.110 | negligible |
|  | *C* | 0.901 | 0.265 | 0.970 | 0.324 | 6.69E-09 | -0.153 | small |
|  | *D* | 0.898 | 0.264 | 0.946 | 0.317 | 1.39E-05 | -0.114 | negligible |
| PM2.5 | *G* | 0.060 | 0.045 | 0.070 | 0.024 | 8.04E-60 | -0.427 | medium |
|  | *F* | 0.058 | 0.023 | 0.086 | 0.109 | 4.54E-49 | -0.386 | medium |
|  | *E* | 0.058 | 0.017 | 0.069 | 0.048 | 2.78E-17 | -0.222 | small |
|  | *A* | 0.049 | 0.013 | 0.023 | 0.013 | 1.41E-217 | 0.825 | large |
|  | *B* | 0.058 | 0.016 | 0.068 | 0.051 | 7.67E-06 | -0.117 | negligible |
|  | *C* | 0.044 | 0.014 | 0.050 | 0.030 | 2.52E-06 | -0.123 | negligible |
|  | *D* | 0.044 | 0.014 | 0.049 | 0.030 | 1.94E-06 | -0.124 | negligible |
| PM4 | *G* | 0.003 | 0.021 | 0.002 | 0.009 | 0.081* | -0.017 | negligible |
|  | *F* | 0.001 | 0.007 | 0.011 | 0.069 | 3.35E-09 | -0.050 | negligible |
|  | *E* | 0.000 | 0.002 | 0.004 | 0.027 | 1.11E-10 | -0.044 | negligible |
|  | *A* | 0.000 | 0.000 | 0.001 | 0.003 | 1.06E-08 | -0.033 | negligible |
|  | *B* | 0.000 | 0.000 | 0.005 | 0.030 | 1.51E-17 | -0.067 | negligible |
|  | *C* | 0.000 | 0.000 | 0.002 | 0.016 | 2.21E-09 | -0.034 | negligible |
|  | *D* | 0.000 | 0.001 | 0.003 | 0.015 | 1.02E-10 | -0.056 | negligible |
| PM10 | *G* | 0.001 | 0.004 | 0.000 | 0.002 | 0.209* | -0.009 | negligible |
|  | *F* | 0.000 | 0.001 | 0.002 | 0.014 | 5.08E-07 | -0.033 | negligible |
|  | *E* | 0.000 | 0.000 | 0.001 | 0.005 | 1.53E-07 | -0.030 | negligible |
|  | *A* | 0.000 | 0.000 | 0.000 | 0.002 | 1.71E-07 | -0.026 | negligible |
|  | *B* | 0.000 | 0.000 | 0.001 | 0.006 | 9.82E-11 | -0.039 | negligible |
|  | *C* | 0.000 | 0.000 | 0.001 | 0.008 | 2.65E-08 | -0.029 | negligible |
|  | *D* | 0.000 | 0.000 | 0.001 | 0.008 | 7.55E-15 | -0.060 | negligible |
| CO2 | *G* | 452.777 | 38.378 | 476.447 | 88.527 | 1.09E-06 | 0.128 | negligible |
|  | *F* | 417.488 | 21.310 | 447.883 | 85.022 | 1.38E-08 | -0.149 | small |
|  | *E* | 465.581 | 24.384 | 462.914 | 63.162 | 4.50E-63 | 0.441 | medium |
|  | *A* | 454.248 | 18.598 | 469.784 | 62.436 | 1.46E-05 | 0.114 | negligible |
|  | *B* | 468.980 | 35.658 | 474.443 | 86.655 | 7.69E-66 | 0.451 | medium |
|  | *C* | 464.864 | 48.653 | 461.177 | 69.010 | 1.14E-64 | 0.447 | medium |
|  | *D* | 406.183 | 44.869 | 403.196 | 84.075 | 2.51E-76 | 0.487 | large |
| VP | All sensors | 1.169 | 0.046 | 1.196 | 0.075 | 4.11E-26 | -0.278 | small |

When the AFU was switched on again at approximately CP2, a dramatic decrease in PM1, PM2.5 and CO_2_ levels was observed throughout the ward space. In order to better understand this event, post hoc decay analysis was performed on the PM1 signal from sensor *A*, which is visualised in Figure 11. From this it can be seen that the PM1 level was approximately constant until about 15:15 when it started to fall for reasons unknown. However, at 15:34 the AFU started up and the rate of decay increased. Analysis of the decay curve using Equation 3 revealed the equivalent ventilation rate achieved by the AFU to be 3.433 air changes per hour (AC/h) in the vicinity of the device. Similar decay rates were also observed in other parts of the ward, as illustrated in Figure 5.

**Figure 11.**
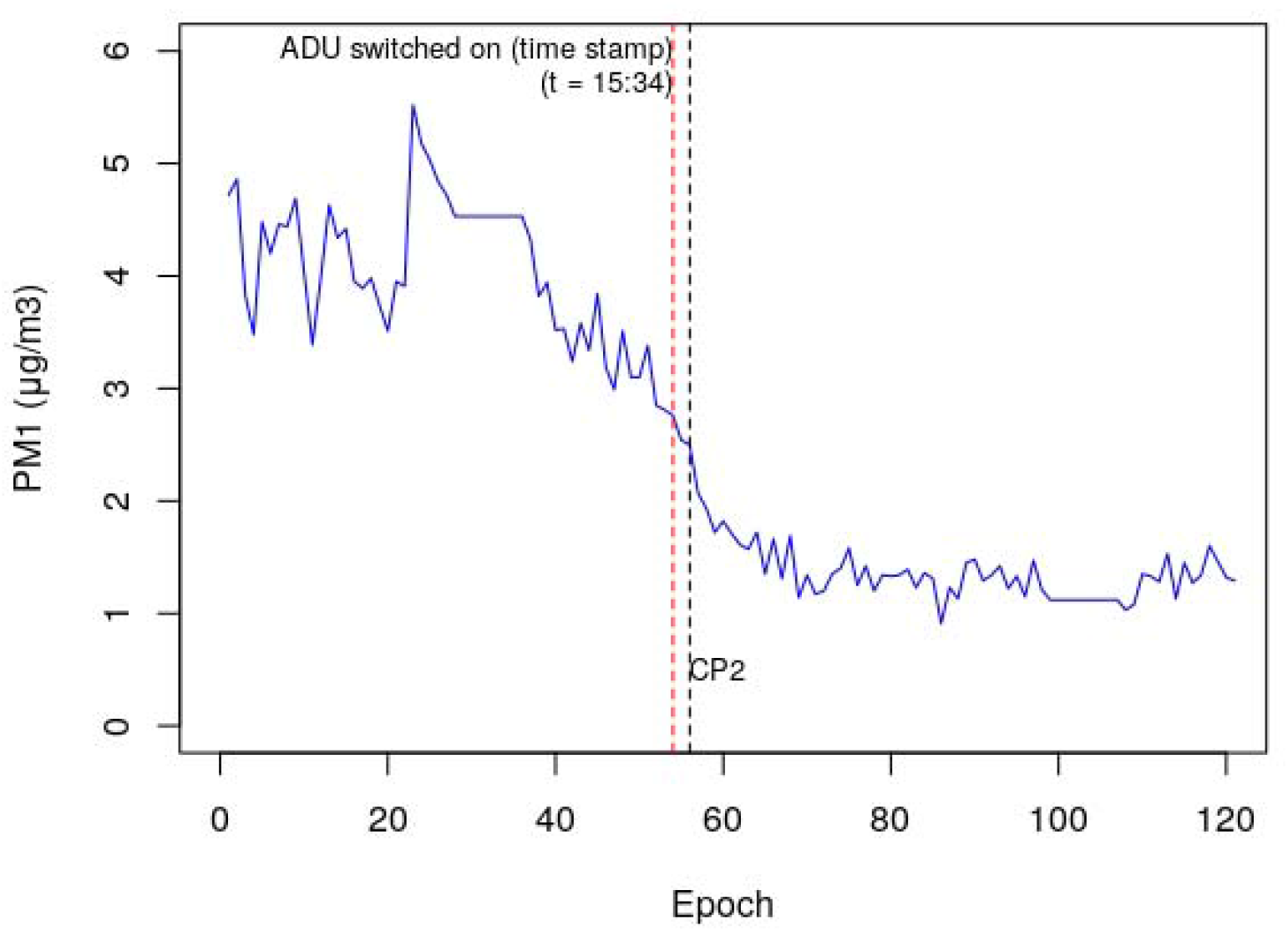
Decay in PM1 levels that occurred at sensor *A* when the AFU was switched on.

## Discussion

This natural experimental study is the first of its kind to comprehensively evaluate both the sequential transport of airborne PM around a medical ward, and to assess the impact of air filtration devices on this. While we did not distinguish between bioaerosols and inert aerosols, the fact that viral particles tend to occur mostly in smaller respiratory aerosols <5 µm diameter [38-40] means that the behaviour of monitored PM signals is likely to have been indicative of any bioaerosols present in the ward air. As such, the study sheds new light on the transport of aerosols around hospital wards that might be helpful in better understanding the dynamics of airborne nosocomial transmission of viruses such as SARS-CoV-2 in clinical settings.

The principal findings of the study were that: (i) the action of the AFU greatly reduced airborne particulate levels of all sizes throughout the ward space; (ii) airborne PM levels of all sizes were positively correlated with indoor CO_2_ levels; and (iii) for all PM sizes, the aerosol particle counts tended to rise and fall simultaneously throughout the ward space, with PM signals from multiple locations being highly correlated. Indoor CO_2_ levels tend to reflect occupancy patterns [41-44], with exhaled CO_2_ being positively correlated with respiratory aerosol emissions [45]. Therefore, the diurnal variation in the PM, CO_2_ and VP signals, together with their close correlation suggests that aerosols associated with human activity were the major driver of the particulate burden. While many of these particulates will be associated with the shedding of skin squamae [46] and activities such as bed making and the washing of patients, etc. [47], respiratory aerosols generated when exhaling or talking are also likely to have made a substantial contribution to the overall burden in the air [14, 16]. Notably, the PM signals indicate that aerosols were widely disseminated around the ward and that they seemed to move freely from sub-compartment to sub-compartment, illustrating the potential for exposure at a distance. The striking effect of the AFU indicates that room air cleaning can be a useful intervention to prevent the build-up of respiratory aerosols and other airborne PM in ward spaces. As such, our findings may help to explain why nosocomial outbreaks of COVID-19, including superspreading events involving multiple sub-compartments, have occurred in many hospital wards despite the application of social distancing measures [19].

One of the most important findings of the study is that airborne particulates rapidly migrated between the various sub-compartments within the ward. This is evident from Table 7, from which it can be seen that strong correlations existed between the PM1 signals from sensors located remotely to each other (e.g. sensors *G* and *A* (PM1: r = 0.834; p<0.001)) during the period when the AFU was not in operation. Similar results were also observed for the PM2.5 signals (e.g. sensors *G* and *A* (PM2.5: r = 0.765; p<0.001)). This indicates that when the AFU was switched off, particulates were being transported considerable distances on air currents around the ward space. This phenomenon may also have been assisted by the turbulent wakes created by the movement of HCWs, which have been shown to transport airborne particulates considerable distances [10, 48]. Collectively this suggests that respiratory aerosols, many of which are <5 µm diameter [9], can be widely disseminated around wards under normal circumstances, and that social distancing measures alone are unlikely to be enough to prevent the transmission of infection [26].

Although the AFU was situated in a single fixed point in the ward, it is notable that its operation resulted in large reductions in PM of all sizes across the ward and not simply proximal to the AFU. This suggests that the strategy of employing a laminar discharge on the AFU with a high velocity to promote good air mixing had the beneficial effect of extending the air cleaning impact of the unit. Indeed, *post hoc* analysis of the decay in the PM1 signal immediately after CP2 (Figure 11) suggests that when the AFU was restarted it provided a total equivalent ventilation rate in the region 3.4 AC/h, an effect that was mirrored throughout the ward space. As the majority of airborne virus is thought to be contained in particles of <5 µm [39, 40, 49], the results reported here imply that appropriately sized and situated air filtration devices, supplementing ward ventilation, may have a significant impact on the nosocomial transmission of viral and other microbial infections. Interestingly, Conway-Morris *et al* [38] observed most viral particles to be in aerosol particles of 1-4 µm diameter, with some in aerosols >4 µm. As well as impacting in the nasopharynx, these smaller aerosols can travel deeper into the lungs to the alveolar level, and there is evidence that this may be associated with more severe disease [12, 13]. Therefore, it would seem that the AFU used in this study was optimally sized for removing viruses from the air, and so it can be hypothesized that the AFU might also reduce the incidence of airborne viral infections on the ward. However, further epidemiological work will be required to confirm this.

Airborne infection risk can be assessed using indoor CO_2_ levels [50], and this has led to the widespread use of CO_2_ monitoring to help mitigate the transmission of SARS-CoV-2 in buildings [30, 31]. Indoor CO_2_ levels can also be used as a surrogate for human activity within buildings [41-44]. With respect to this, we found that when the AFU was not in operation there was a moderately strong positive correlation between PM and CO_2_, especially for the smaller particulates, and also with VP - although the correlations involving VP exhibited considerable variability. Furthermore, both CO_2_ and VP exhibited diurnal variation that was likely related to human activity and exhaled breath. Collectively, this suggests that most of the particulates found in the ward air were associated with human activity, and as such this corroborates Roberts *et al* [47] finding that aerosol production on a respiratory ward was associated with activities such as bed making, ward rounds, curtain drawing, etc. all of which liberated copious quantities of PM into the air.

Interestingly, when the AFU was in operation, both the PM counts and the CO_2_ levels were greatly reduced, which was an unexpected finding because the AFU should have had no direct effect on CO_2_ levels within the ward. While the reasons for this observation are unclear, it may be that the AFU promoted better mixing of the air on the ward, preventing the CO_2_ from stratifying, and thus reducing its concentration in the lower part of the room space. Alternatively, the AFU may also have increased air velocities within the ward space to such an extent that additional ‘fresh air’ might have been entrained in from outside. While both explanations appear plausible, it is difficult to explain the magnitude to the reduction in CO_2_ levels observed in Figure 9 simply by better mixing alone, especially as the sensors were mounted >1.5 m above floor level. It might therefore be that the low PM levels observed on the ward while the AFU was in operation arose from a combination of the air filtration and improved ventilation of the space. However, further investigation will be required to determine whether or not this was the case.

With regard to the change in the AFU fan speed that occurred on the morning of the 4^th^ August, the results of our *post hoc* analysis were somewhat inconclusive. Although increasing the fan speed from setting 2 to 3 appeared only to have a localised effect in the vicinity of sensor *A*, we cannot say for certain that this is the case. This is because the change in the speed setting approximately coincided with the time (9:00 am) when activity on the ward greatly increased. Thus even though the PM levels observed for most sensors actually increased when the fan speed was increased, we cannot be confident that increasing the fan speed had no beneficial effect. It could be the case that due to increased activity, the PM counts throughout the ward might have been much higher, and that increasing the fan speed actually limited the magnitude of this potential rise.

Although the positive correlations exhibited between the PM, CO_2_ and VP signals indicate that the liberation of PM into the air was strongly associated with human activity on the ward (e.g. talking, ward rounds, bed making, etc.) as others have found [47, 51], we were not able to determine the proportion of airborne particulates that comprised respiratory aerosols. Previous studies have shown that respiratory viruses are most likely to be recovered from particles <5 µm [38-40, 49], suggesting that they are contained in exhaled bioaerosols that have undergone rapid evaporation [7]. Such aerosols are thought to reduce in size to approximately 30% of their original diameter under normal room conditions [52], which means that exhaled respiratory droplets as large as 100 µm diameter have the potential to transmit SARS-CoV-2 because of evaporation [6, 53]. Indeed it has been shown that when speaking, 80-94% of the respiratory droplets produced are 100 µm or less [54], indicating that the vast majority of the droplets exhaled become aerosolised, with only droplets larger than this threshold behaving ballistically [55, 56]. Having said this, because aerosol particles >10 µm diameter tend to fall out of the air fairly rapidly within a few minutes, it means that the ones most likely to persist in room air will be the smaller aerosols, say <5 µm, which may explain why viruses are most likely to be recovered from aerosols in this size range [38-40, 49]. Given that median (range) aerosol particle emission rates of: 135 (85-691) particles/s for breathing; 270 (120-1380) particles/s for normal talking; and 570 (180-1760) particles/s for loud talking, have been recorded [16], this suggest that on a typical medical ward many thousands of respiratory aerosols are likely to liberated into the air when the ward is busy, with many remaining airborne for some considerable time [54].

Although the study was a natural experiment that came about because of a chance mistake, the data yielded has been highly informative, highlighting the ease with which respiratory sized aerosols can be widely disseminated around medical wards - implying that social distancing measures alone are unlikely to be enough to prevent the nosocomial spread of viral infections such as SARS-CoV-2. The data also indicate that air filtration technologies have considerable potential to reduce transmission of airborne pathogens in shared spaces on the basis that they may reduce the viral dose inhaled and thus the likelihood of disease transmission. Air filtration devices may therefore be applicable not only to pathogens traditionally considered as airborne such as measles and tuberculosis (TB), but also to fungal and bacterial infections which may have a component of aerial dissemination in the transmission cycle such as *C. diff*. spores [57].

While the study yielded useful insights, it is important to remember its limitations. For example, while historical data existed concerning air change rates on the ward, because it was a natural experiment we do not know the actual baseline ventilation rates that occurred on the 3^rd^ and 4^th^ August 2021. Furthermore, although we generated detailed data regarding airborne particulates, CO_2_ and VP we did not record staff occupancy and movement, or window and door opening. As a result, our conclusions around the mechanisms by which these particulates and gases were generated, and the mechanism by which the AFU altered them remain hypothesis generating rather than confirmed. However, given the known nature of microbial bioaerosols in hospitals [9, 38-40], it is highly likely that reduction in these by any mechanism will improve the quality of indoor air and reduce infection transmission. Also, the study was conducted in a single ward in a hospital built before the regulations required higher air change rates and the installation of doors to separate bays, and this is likely to have influenced the free flow of particulates between the sub-compartments in the ward. As such, we cannot be certain that the results will be generalisable to other settings, although wards of this age and design are not uncommon within the UK or indeed other settings worldwide.

It is worth noting that the study reported here only came about because of a mistake that occurred when the AFU was being commissioned as part of a larger study involving both intervention and control wards. As such, it highlights the need to use PM and CO_2_ from sensors to commission such devices. Without these, it is not possible to demonstrate that the filtration device is having an effect, let alone measure the magnitude of that effect. Furthermore, the study also highlighted the need to ensure that any filtration units used have a long-range impact, rather than just a localised effect. Unfortunately, air filtration devices are all too often placed in rooms without any consideration given to commissioning or indeed how their performance can be validated. It is therefore recommended that future work be undertaken to identify how room air filtration devices should be best be applied, optimised and validated in order maximise their effectiveness in the clinical setting.

## Conclusions

This study builds on previous work showing that air filtration units utilising HEPA filters and UVC light have the potential ability to reduce microbial contamination in ward air [38]. As such, it demonstrates that a single high throughput (2550-3000 m^3^/h) AFU can greatly reduce the burden of particulate matter in air throughout a large hospital ward. The application of a combined HEPA and UV-C AFU in a medicine for older people ward led to substantial reductions in airborne PM levels, most notably in the range of particle sizes that are associated with infectious respiratory viruses, such as SARS-CoV-2. The study also found that airborne particulates associated with human activity were able to freely migrate considerable distances throughout the ward space, an observation that suggests that social distancing measures alone are unlikely to be enough to prevent the transmission of viral infection. Collectively, this suggests that the application of appropriately sized air filtration units in poorly ventilated hospital wards has the potential to significantly reduce rates of nosocomial viral infection in such settings and warrants further investigation. Furthermore, the study highlights the importance of commissioning such devices, considering their effect on air flow and the removal of contaminants in order to optimise the systems ability to clean the air in the ward space.

## Data Availability

All data produced in the present study are available upon reasonable request to the authors

## Supplementary Material

**Table S1.** Descriptive statistics for the various sensor signals divided into change point blocks.

| Signal | Sensor ID | Before CP1<br>Mean | Before CP1<br>SD | CP1-CP2<br>Mean | CP1-CP2<br>SD | CP2-CP3<br>Mean | CP2-CP3<br>SD | CP3-CP4<br>Mean | CP3-CP4<br>SD | CP4-CP5<br>Mean | CP4-CP5<br>SD | After CP5<br>Mean | After CP5<br>SD |
| --- | --- | --- | --- | --- | --- | --- | --- | --- | --- | --- | --- | --- | --- |
| PM1 | *G* | 2.673 | 0.339 | 3.728 | 1.299 | 1.424 | 0.683 | 0.906 | 0.205 | 1.054 | 0.424 | 1.285 | 0.189 |
|  | *F* | 2.401 | 0.320 | 5.054 | 2.347 | 1.366 | 0.359 | 0.935 | 0.232 | 1.134 | 0.727 | 1.370 | 0.204 |
|  | *E* | 2.273 | 0.303 | 2.827 | 0.696 | 1.364 | 0.253 | 0.947 | 0.226 | 0.898 | 0.412 | 1.317 | 0.203 |
|  | *A* | 0.830 | 0.178 | 2.961 | 0.871 | 1.361 | 0.213 | 0.939 | 0.221 | 0.291 | 0.140 | 0.614 | 0.197 |
|  | *B* | 2.597 | 0.323 | 2.607 | 0.572 | 1.373 | 0.257 | 0.950 | 0.215 | 0.894 | 0.449 | 1.223 | 0.164 |
|  | *C* | 2.303 | 0.283 | 2.599 | 0.574 | 1.284 | 0.319 | 0.833 | 0.186 | 0.822 | 0.393 | 1.094 | 0.173 |
|  | *D* | 2.221 | 0.257 | 2.641 | 0.613 | 1.260 | 0.339 | 0.834 | 0.186 | 0.826 | 0.398 | 1.048 | 0.171 |
|  | Mean | **2.185** | **0.286** | **3.202** | **0.996** | **1.347** | **0.346** | **0.906** | **0.210** | **0.846** | **0.420** | **1.136** | **0.186** |
| PM2.5 | *G* | 0.153 | 0.020 | 0.394 | 0.308 | 0.102 | 0.103 | 0.052 | 0.012 | 0.065 | 0.033 | 0.074 | 0.012 |
|  | *F* | 0.138 | 0.020 | 0.747 | 0.650 | 0.082 | 0.042 | 0.054 | 0.014 | 0.094 | 0.160 | 0.079 | 0.012 |
|  | *E* | 0.131 | 0.018 | 0.217 | 0.125 | 0.079 | 0.019 | 0.054 | 0.013 | 0.062 | 0.069 | 0.076 | 0.013 |
|  | *A* | 0.040 | 0.009 | 0.201 | 0.140 | 0.066 | 0.011 | 0.046 | 0.011 | 0.016 | 0.011 | 0.030 | 0.010 |
|  | *B* | 0.149 | 0.019 | 0.198 | 0.106 | 0.080 | 0.016 | 0.055 | 0.013 | 0.064 | 0.074 | 0.070 | 0.010 |
|  | *C* | 0.115 | 0.044 | 0.153 | 0.078 | 0.062 | 0.017 | 0.041 | 0.010 | 0.045 | 0.043 | 0.053 | 0.009 |
|  | *D* | 0.108 | 0.013 | 0.168 | 0.094 | 0.062 | 0.018 | 0.041 | 0.010 | 0.047 | 0.042 | 0.051 | 0.010 |
|  | Mean | **0.119** | **0.020** | **0.297** | **0.214** | **0.076** | **0.032** | **0.049** | **0.012** | **0.056** | **0.062** | **0.062** | **0.011** |
| PM4 | *G* | 0.000 | 0.000 | 0.146 | 0.194 | 0.017 | 0.053 | 0.000 | 0.001 | 0.004 | 0.013 | 0.000 | 0.000 |
|  | *F* | 0.000 | 0.004 | 0.369 | 0.425 | 0.004 | 0.018 | 0.000 | 0.000 | 0.024 | 0.100 | 0.000 | 0.000 |
|  | *E* | 0.000 | 0.000 | 0.044 | 0.073 | 0.001 | 0.005 | 0.000 | 0.000 | 0.008 | 0.040 | 0.000 | 0.000 |
|  | *A* | 0.000 | 0.000 | 0.046 | 0.085 | 0.000 | 0.000 | 0.000 | 0.000 | 0.001 | 0.005 | 0.000 | 0.000 |
|  | *B* | 0.000 | 0.000 | 0.039 | 0.066 | 0.000 | 0.000 | 0.000 | 0.000 | 0.010 | 0.044 | 0.000 | 0.000 |
|  | *C* | 0.002 | 0.035 | 0.021 | 0.047 | 0.000 | 0.000 | 0.000 | 0.000 | 0.005 | 0.024 | 0.000 | 0.000 |
|  | *D* | 0.000 | 0.002 | 0.031 | 0.058 | 0.001 | 0.003 | 0.000 | 0.000 | 0.006 | 0.021 | 0.000 | 0.000 |
|  | Mean | **0.000** | **0.006** | **0.099** | **0.135** | **0.003** | **0.011** | **0.000** | **0.000** | **0.008** | **0.035** | **0.000** | **0.000** |
| PM10 | *G* | 0.000 | 0.000 | 0.030 | 0.039 | 0.003 | 0.011 | 0.000 | 0.000 | 0.001 | 0.003 | 0.000 | 0.000 |
|  | *F* | 0.000 | 0.001 | 0.074 | 0.086 | 0.001 | 0.003 | 0.000 | 0.000 | 0.005 | 0.020 | 0.000 | 0.000 |
|  | *E* | 0.000 | 0.000 | 0.009 | 0.015 | 0.000 | 0.001 | 0.000 | 0.000 | 0.002 | 0.008 | 0.000 | 0.000 |
|  | *A* | 0.000 | 0.000 | 0.024 | 0.044 | 0.000 | 0.000 | 0.000 | 0.000 | 0.001 | 0.003 | 0.000 | 0.000 |
|  | *B* | 0.000 | 0.000 | 0.007 | 0.013 | 0.000 | 0.000 | 0.000 | 0.000 | 0.002 | 0.009 | 0.000 | 0.000 |
|  | *C* | 0.001 | 0.018 | 0.011 | 0.025 | 0.000 | 0.000 | 0.000 | 0.000 | 0.002 | 0.012 | 0.000 | 0.000 |
|  | *D* | 0.000 | 0.001 | 0.017 | 0.031 | 0.000 | 0.001 | 0.000 | 0.000 | 0.003 | 0.011 | 0.000 | 0.000 |
|  | Mean | **0.000** | **0.003** | **0.025** | **0.036** | **0.001** | **0.002** | **0.000** | **0.000** | **0.002** | **0.009** | **0.000** | **0.000** |
| CO2 | *G* | 469.307 | 13.578 | 815.175 | 139.885 | 489.345 | 84.741 | 446.327 | 13.966 | 518.482 | 112.203 | 441.099 | 33.428 |
|  | *F* | 445.665 | 8.453 | 734.244 | 80.023 | 433.748 | 47.122 | 414.619 | 9.472 | 485.906 | 113.827 | 415.909 | 13.646 |
|  | *E* | 470.582 | 8.077 | 691.243 | 72.783 | 481.153 | 52.930 | 462.834 | 12.553 | 503.166 | 74.680 | 429.066 | 12.241 |
|  | *A* | 467.185 | 10.276 | 713.817 | 80.280 | 464.744 | 40.040 | 452.397 | 10.138 | 502.820 | 79.962 | 442.005 | 10.755 |
|  | *B* | 466.868 | 10.582 | 818.856 | 115.035 | 500.493 | 73.982 | 463.421 | 18.145 | 523.991 | 108.488 | 432.777 | 11.614 |
|  | *C* | 443.028 | 22.364 | 833.165 | 121.853 | 516.107 | 102.240 | 455.824 | 20.135 | 497.942 | 87.750 | 430.261 | 14.356 |
|  | *D* | 398.425 | 8.607 | 748.612 | 113.939 | 451.045 | 91.525 | 398.269 | 21.924 | 450.621 | 105.655 | 363.316 | 12.350 |
|  | Mean | **451.580** | **11.705** | **765.016** | **103.400** | **476.662** | **70.369** | **441.956** | **15.190** | **497.561** | **97.509** | **422.062** | **15.484** |
| VP | All sensors | 1.296 | 0.035 | 1.368 | 0.051 | 1.210 | 0.027 | 1.162 | 0.045 | 1.161 | 0.056 | 1.225 | 0.076 |

Legend: PM1 – Particulate matter (PM) fraction < 1 µm diameter (µg/m<sup>3</sup>); PM2.5 – PM fraction between 1 and 2.5 µm diameter (µg/m<sup>3</sup>); PM4 – PM fraction between 2.5 and 4 µm diameter (µg/m<sup>3</sup>); PM10 – PM fraction between 4 and 10 µm diameter (µg/m<sup>3</sup>); CO2 – carbon dioxide (ppm); VP – vapour pressure (kPa); and CP1-5 – change points 1 to 5.

## Notes

**Ethics, funding and permissions:** The work presented in this manuscript did not constitute research under the UK Policy Framework for Health and Social Care Research and was not subject to formal ethical review. Subsequently, this work led to a larger project which is ongoing, the Addenbrooke’s Air Disinfection Study (AAirDS, Chief Investigator: Dr Victoria L Keevil). AAirDS is funded by the UK Health Security Agency and was approved by the NHS Health Research Authority and Health and Care Research Wales (HCRW) (IRAS ID 299336) and the Central Bristol Research Ethics Committee (REC reference: 22/SW/0010). Dr Victoria L Keevil is funded by a MRC/NIHR Clinical Academic Research Partnership Grant (CARP; grant code: MR/T023902/1). Dr Andrew Conway Morris is supported by a Clinician Scientist Fellowship from the Medical Research Council (MR/V006118/1).

### Competing Interest Statement

Darren Sloof is the founder, director and shareholder of Air Purity Ltd. Air Purity designed and supplied the air filtration units and air sensors used in this study. Air Purity Ltd had no role in the study design or analysis of the data. Darren Sloof did however collect the data and liaise with other authors over its interpretation.
Dr Andrew Conway Morris is a member of the scientific advisory board of Cambridge Infection Diagnostics.
Dr Theodore Gouliouris has received materials from Shionogi for conducting a laboratory evaluation.
The other authors have no competing interests.

### Funding Statement

The work presented in this manuscript was a preliminary study which subsequently led to a larger project which is ongoing, the Addenbrooke's Air Disinfection Study (AAirDS, Chief Investigator: Dr Victoria L Keevil). AAirDS is funded by the UK Health Security Agency and was approved by the NHS Health Research Authority and Health and Care Research Wales (HCRW) (IRAS ID 299336) and the Central Bristol Research Ethics Committee (REC reference: 22/SW/0010).
Dr Victoria L Keevil is funded by a MRC/NIHR Clinical Academic Research Partnership Grant (CARP; grant code: MR/T023902/1).
Dr Andrew Conway Morris is supported by a Clinician Scientist Fellowship from the Medical Research Council (MR/V006118/1).
Air Purity Ltd. designed and supplied the air filtration units and air sensors used in this study. Air Purity Ltd had no role in the study design or analysis of the data. Darren Sloof, the founder, director and shareholder of Air Purity did however collect the data and liaise with other authors over its interpretation.

